# Associations between brain imaging and polygenic scores of mental health and educational attainment in children aged 9-11

**DOI:** 10.1101/2022.02.01.22270003

**Authors:** Sara Fernandez-Cabello, Dag Alnæs, Dennis van der Meer, Andreas Dahl, Madelene Holm, Rikka Kjelkenes, Ivan I. Maximov, Linn B. Norbom, Mads L. Pedersen, Irene Voldsbekk, Ole A. Andreassen, Lars T. Westlye

## Abstract

Psychiatric disorders are highly heritable and polygenic, and many have their peak onset in late childhood and adolescence, a period of tremendous changes. Although the neurodevelopmental antecedents of mental illness are widely acknowledged, research in youth population cohorts is still scarce, preventing our progress towards the early characterization of these disorders. We included 7,124 children (9-11 years old) from the Adolescent Brain and Cognitive Development Study to map the associations of structural and diffusion brain imaging with common genetic variants and polygenic scores for psychiatric disorders and educational attainment. We used principal component analysis to derive imaging components, and calculated their heritability. We then assessed the relationship of imaging components with genetic and clinical psychiatric risk with univariate models and Canonical correlation analysis (CCA). Most imaging components had moderate heritability. Univariate models showed limited evidence and small associations of polygenic scores with brain structure at this age. CCA revealed two significant modes of covariation. The first mode linked higher polygenic scores for educational attainment with less externalizing problems and larger surface area. The second mode related higher polygenic scores for schizophrenia, bipolar disorder, and autism spectrum disorder to higher global cortical thickness, smaller white matter volumes of the fornix and cingulum, larger medial occipital surface area and smaller surface area of lateral and medial temporal regions. While cross-validation suggested limited generalizability, our results highlight the potential of multivariate models to better understand the transdiagnostic and distributed relationships between mental health and brain structure in late childhood.

## 1. Introduction

Under the influence of genetic and environmental factors, human brain development covers a protracted period of time from prenatal life to early adulthood. Departures from brain developmental trajectories have been suggested to be fundamental to the emergence of psychiatric disorders and may be observed long before symptoms begin (Birnbaum and Weinberger, 2017; Insel, 2010; Thapar and Riglin, 2020; Weinberger, 1987). Many psychiatric disorders have their peak onset during late childhood and adolescence (Solmi et al., 2021), a period that coincides with tremendous biological, cognitive and behavioral changes and exposures (Dahl et al., 2018; Larsen and Luna, 2018; Marco et al., 2011). Psychiatric disorders are highly heritable and polygenic. With the growing availability of summary statistics from genome-wide association studies (GWAS) and genotyping data, polygenic scores (PRS) can be used to study inter-individual differences in the genetic disposition to psychiatric disorders and other traits. Understanding the impact of the genetic determinants of disease on the brain during important developmental periods may foster our knowledge of disease pathophysiology and progression. Thus, the central aim of this study is to provide a more complete picture of the genetic effects on multimodal brain imaging metrics in a large population sample of children and adolescents.

Available samples with thousands of individuals deeply genotyped and phenotyped have recently offered new opportunities to investigate the impact of polygenic risk for psychiatric disorders in the general population. If psychiatric disorders are not distinct entities but represent the ends of developmental continuums (Cuthbert, 2014; Weinberger, 2017), then studying the full range of variation of risk in the general population may provide major insights into disease processes (Cuthbert, 2014; Weinberger, 2017, 1987). Prior studies have examined the relationship of PRS of psychiatric disorders and the brain in adult population samples. For example, individuals with high PRS of schizophrenia show similar anatomical patterns as those observed in patients (Alnæs et al., 2019; van Erp et al., 2018), suggesting that PRS in asymptomatic individuals may convey sensitive information to disease-related processes (Westlye et al., 2019). Whether brain differences are already observable at earlier stages of development is an essential question that remains to be fully examined. Research in younger population samples is at present scarce and current small sample sizes may hinder the power to detect small effects. Nonetheless, recent studies point to cortical differences in individuals with high PRS already observed in youth (Khundrakpam et al., 2020; Kirschner et al., 2021).

Psychiatric disorders are highly polygenic and are thus likely to have distributed effects on the brain (Birnbaum and Weinberger, 2017; Insel, 2010; Ripke et al., 2020), possibly observed in multiple neuroimaging modalities that are sensitive to different tissue properties (Lazari et al., 2021; Natu et al., 2019; Ripke et al., 2020; Rokicki et al., 2021). Besides, brain maturation during late childhood and adolescence is highly spatially heterogeneous and dynamic, covering increases in surface area, apparent cortical thinning or white matter microstructural changes that possibly reflect synaptic remodeling and myelination (Norbom et al., 2021; Ritchie et al., 2019; Westlye et al., 2010; Zhou et al., 2015). Multimodal neuroimaging can aid the mechanistic interpretation of neuroimaging results (Natu et al., 2019) and increase certainty and sensitivity to detect associations between brain imaging and other traits. Furthermore, there is growing recognition of the distributed nature of the brain’s organizational principles. Rather than independent and discrete, brain regions and pathways seem to follow organizational axes likely reflecting functional hierarchies (Colby et al., 2011; Du and Buckner, 2021; Huntenburg et al., 2018; Sotiras et al., 2017; Sydnor et al., 2021; Valk et al., 2020). Compared to treating all brain regions or tracts independently in mass-univariate analyses, multivariate modeling techniques that capture the covariance patterns of multimodal brain imaging and other variables appear more biologically meaningful. Benefits notwithstanding, multivariate models are prone to overfitting, and statistical significance does not guarantee generalizability even in large samples (McIntosh, 2021; Smith and Nichols, 2018). Both careful statistical significance testing and generalizability should be addressed.

Taken together, with the main aim of investigating the associations between brain imaging and genetic risk for psychiatric disorders in late childhood we here relied on 1) Youth population samples, 2) inter-individual differences in polygenic scores for mental disorders and educational attainment, 3) multimodal brain imaging measures and 4) multivariate models with careful cross-validation. We capitalized on the multi-site US-based Adolescent Brain and Cognitive Development (ABCD) Study, that has recruited ∼11,800 children aged 9-11 years at baseline (Casey et al., 2018). Multiple macro and microstructural imaging data were generated using standardized pipelines and decomposed into their axes of maximal variation (“imaging components”) spanning global and regional components. Using single nucleotide polymorphism (SNP) data from all individuals, we investigated the heritability – the proportion of the brain imaging components variance explained by differences in SNPs – through Genomic Complex Trait Analysis (GCTA). We then tested for associations between polygenic scores for various psychiatric disorders and educational attainment and the imaging components as well as clinical symptomatology scores of externalizing and internalizing behaviors. Finally, we investigated the modes of variation between brain imaging components and polygenic scores and clinical risk with canonical correlation analysis (CCA), addressing both statistical significance and generalizability.

Following past reports, we hypothesized that all imaging measures would show moderate to high heritability (L. T. Elliott et al., 2018; Grasby et al., 2020; Li et al., 2018; Schmitt et al., 2020; Strike et al., 2019) and expected small and widespread associations between PRS of psychiatric disorders and brain imaging components (Alnæs et al., 2019; Kirschner et al., 2021; Westlye et al., 2019). As a result, we here attempt to provide a more complete picture of the genetic effects on brain structure during this important developmental period.

## 2. Methods and Materials

### 2.1 Sample

We used data from the ABCD 3.0 release (DOI: http://dx.doi.org/10.15154/1519007). The final sample of the present study after quality control (QC; see below) included 7,124 individuals (3,388 female/3,736 male) with a mean age of 9.92 years (standard deviation=0.62).

### 2.2 Ethical Approval

All procedures were approved by a central Institutional Review Board (IRB) at the University of California, San Diego, and in some cases by individual site IRBs (e.g. Washington University in St. Louis). Parents or guardians provided written informed consent after the procedures had been fully explained and children assented before participation in the study.

### 2.3 MRI data

Structural T1-weighted and diffusion-weighted measures from bilateral regions of interest (ROIs) were included from the tabulated 3.0 imaging release baseline data. The imaging processing pipelines are explained in detail elsewhere (Hagler et al., 2019). Briefly, structural imaging data were corrected for gradient nonlinearity distortions and intensity inhomogeneities and were rigidly registered to an in-house average adult template (Hagler et al., 2019). The images were then submitted to FreeSurfer v5.3 to obtain averages of cortical thickness and surface area within 68 bilateral cortical ROIs (n total cortical ROIs = 136) from the Desikan-Killiany parcellation (Desikan et al., 2006). Diffusion-weighted data were corrected for eddy currents, signal dropouts, head motion, gradient nonlinearities and B0 distortions and were registered to the T1w images and resampled to a standard orientation. 37 major white matter tracts were labeled using a probabilistic atlas – AtlasTrack; Hagler et al., 2009) – containing prior information of the tracts excluding voxels containing primarily gray matter or cerebrospinal fluid (see Hagler et al., 2019 supplementary table 6 for a complete list of the tracts). Average values within the tracts of several microstructural metrics were obtained after estimating diffusion tensor imaging (DTI) and restriction spectrum imaging (RSI; White et al., 2013) models. From the DTI models, metrics that quantify different aspects of water diffusivity included fractional anisotropy (DTI-FA; magnitude of directional (spherical) diffusion), mean diffusivity (DTI-MD; total amount of diffusion), longitudinal diffusivity (DTI-LD; diffusion parallel to tracts) and transverse diffusivity (DTI-TD; diffusion perpendicular to tracts). From the RSI models, averaged measures within the tracts included normalized directional restricted diffusion (RSI-Res) and normalized directional hindered diffusion (RSI-Hind), presumably reflecting oriented diffusion of the intracellular and extracellular components respectively. In addition, the volume of the white matter tracts in mm^3 was included (DTI-Vol; (n total white matter ROIs = 259). See Supplementary Table 1. The QC of the tabulated magnetic resonance imaging (MRI) data included: 1) exclusion of participants that did not pass the ABCD recommended QC (imgincl_t1w_include==0 & imgincl_dmri_include==0). The recommended QC was based on the quality of the raw data (e.g., discard images with severe artifacts or poor quality) and postprocessing quality (e.g., manual QC of surface reconstruction or accuracy of fiber tract segmentation, automated QC measures); 2) set extreme values (robust median-based z-scores> 3) to missing values; 4) exclusion of participants with more than 10% of missing values in each imaging modality (structural and diffusion MRI) and 5) imputation of the remaining missing values with k-nearest neighbors algorithm (k=3). 8,806 participants remained after these preprocessing steps. Visual assessments of the imputation quality are provided in the Supplementary information and show similar distributions between the observed an imputed data. However, these descriptive checks do not rule out the possibility that the distributions are significantly different. So in order to ensure that the imputation method did not affect the main results, the multivariate CCA analysis was repeated without excluding and imputing extreme observations (n=9,071).

**Table 1.** MRI PCA results. A PCA was performed with the ROI data of each imaging modality and the first three principal components were retained. The table shows the proportion of the total variance and the cumulative proportion of the variance explained by the first three principal components of each modality. For the full loadings please see Supplementary Table 2.

| Modality | PC | Variance explained | Cummulative variance explained |
| --- | --- | --- | --- |
| Cortical Thickness | PC1 | 0.318 | 0.318 |
| Cortical Thickness | PC2 | 0.045 | 0.363 |
| Cortical Thickness | PC3 | 0.035 | 0.398 |
| Surface Area | PC1 | 0.350 | 0.350 |
| Surface Area | PC2 | 0.042 | 0.391 |
| Surface Area | PC3 | 0.032 | 0.423 |
| DTI Vol | PC1 | 0.609 | 0.609 |
| DTI Vol | PC2 | 0.049 | 0.658 |
| DTI Vol | PC3 | 0.043 | 0.701 |
| DTI FA | PC1 | 0.358 | 0.358 |
| DTI FA | PC2 | 0.099 | 0.457 |
| DTI FA | PC3 | 0.069 | 0.526 |
| DTI MD | PC1 | 0.571 | 0.571 |
| DTI MD | PC2 | 0.087 | 0.658 |
| DTI MD | PC3 | 0.047 | 0.705 |
| DTI LD | PC1 | 0.352 | 0.352 |
| DTI LD | PC2 | 0.093 | 0.445 |
| DTI LD | PC3 | 0.068 | 0.513 |
| DTI TD | PC1 | 0.501 | 0.501 |
| DTI TD | PC2 | 0.088 | 0.590 |
| DTI TD | PC3 | 0.057 | 0.646 |
| RSI-Res | PC1 | 0.455 | 0.455 |
| RSI-Res | PC2 | 0.084 | 0.539 |
| RSI-Res | PC3 | 0.070 | 0.609 |
| RSI-Hind | PC1 | 0.319 | 0.319 |
| RSI-Hind | PC2 | 0.096 | 0.415 |
| RSI-Hind | PC3 | 0.075 | 0.490 |

### 2.4 Genetic data

The genotyped data were obtained using an Affymetrix NIDA SmokeScreen array containing 733,293 variants (Baurley et al., 2016). QC was performed based on calling signals and variant rate calls, following previous recommendations (Lam et al., 2019). See NIMH experiment #1194 for more information. Imputation was performed with the multi-ethnic TOPMed reference panel. Post-imputation QC was performed using plink (Purcell et al., 2007); http://pngu.mgh.harvard.edu/purcell/plink/) included the following steps: exclusion of low imputation scores and duplicates, genotype and individual missingness (--geno 0.1 --mind 0.1), filter out low minor allele frequencies (--maf 0.05), and deviation from Hardy-Weinberg equilibrium (--hwe 1e-6). After these steps, 4.606,001 variants and 11,101 individuals remained. Data from the genotyping plate 461 were excluded (n=82) following the recommendations. Regions in high linkage-disequilibrium including the major-histocompatibility complex were excluded (https://genome.sph.umich.edu/wiki/Regions_of_high_linkage_disequilibrium_(LD). The ABCD cohort includes both unrelated and related individuals (siblings, twins, etc.). Due to possible confounds of the family structure, we calculated pairwise family relationships from the genotyped data (ABCD_release_3.0_QCed) using King v. 2.24 (http://people.virginia.edu/~wc9c/KING/manual.html), and only one individual from a family of twins and first siblings was kept for further analyses, with the reasoning that this step will exclude most family relations likely to share close environments without substantially reducing the sample size. After merging these data with the valid MRI data after QC, 7,124 individuals remained. As population structure can be a confound due to the presence of different alleles in different populations, in addition to accounting for population ancestry components (see below), we repeated the main analyses in participants with European genetic ancestry (*genetic_af_european* > 0.90; n=3,841).

### 2.5 Polygenic scores

Polygenic scores are weighted sums of risk alleles weighted by their effect sizes as found through previous GWAS. To study the relationships between PRS of psychiatric disorders and brain imaging phenotypes, we constructed PRS for various disorders spanning neurodevelopmental and later onset psychiatric disorders, making use of the latest and largest publicly available GWAS summary statistics: Attention-Deficit/Hyperactivity Disorder (PRS-ADHD, Demontis et al., 2019), Autism Spectrum Disorder (PRS-ASD, The Autism Spectrum Disorders Working Group of The Psychiatric Genomics Consortium, 2017), Bipolar Disorder (PRS-BIP, Stahl et al., 2019), Major Depressive Disorder (PRS-MDD, Wray et al., 2018), Obsessive-Compulsive Disorder (PRS-OCD, International Obsessive Compulsive Disorder Foundation Genetics Collaborative (IOCDF-GC) and OCD Collaborative Genetics Association Studies (OCGAS), 2018), Schizophrenia (PRS-SCZ, Trubetskoy et al., 2022) and a Cross-Disorder PRS (PRS-Multiple, Lee et al., 2019) capturing pleiotropy between multiple diagnoses including anorexia nervosa, ADHD, ASD, BIP, MDD, OCD SCZ, and Tourette syndrome. In addition, we included a PRS of Educational attainment (PRS-Edu, Lee et al., 2018) given its possible implication in brain development and higher prediction accuracy. All PRS was created using PRSice-2 (Choi and O’Reilly, 2019). Because SNPs are in linkage disequilibrium, a clumping procedure to retain independent SNPs was used with default settings that included clumping any SNP within a 250kb window and a correlation of 0.1. PRS scores for each individual were calculated over a range of p-value thresholds (min=5e-8, max=0.5, interval=5e-05).

### 2.6 Dimensional symptoms and height

In addition to exploring the relationship between PRS and brain imaging, we further included dimensional clinical scores to compare their relationship with imaging measures. We included dimensional symptomatology scores from the parent-reported Child-Behavior checklist questionnaire (CBCL), which assesses a wide range of behavioral and emotional problems in the past six months. These problems can be broadly categorized as internalizing, covering a range of emotional problems (mood, anxiety and depression, etc.) or externalizing, indicating overt behavioral conflicts and violation of social norms. Internalizing and externalizing problems were derived from the following syndromic scales: anxious/depressed, withdrawn/depressed and somatic complaints, social problems, thought problems, rule-breaking behavior and aggressive behavior. We obtained the raw scores for externalizing and internalizing behaviors. We computed robust z-scores based on each score’s deviation from the median absolute deviation (MAD). Scores with a robust-z >4 (4 x MAD) were considered extreme and were imputed with the R package *mice*. Less than 10% of values were imputed in each variable. In addition, we included a measure of anthropometric height to compare the genetic effects on brain imaging measures and height. The same QC steps were performed as for the CBCL variables. The results were reassessed without imputing the extreme values. All variables were inverse-ranked normally transformed and z-scored before analyses.

### 2.7 Principal Component Analysis (PCA)

PCA seeks to reduce the dimensionality of a dataset by transforming the initial variables into a smaller set of orthogonal axes. We used PCA on both the MRI ROI measures and the PRS scores to downscale the data into the axes that captured the largest amount of variation in the data. The PRS scores were calculated over a range of thresholds. Since the optimal *p-value* threshold for the PRS is unknown, using PCA to reduce the PRS scores provides unbiased estimates for association tests while maximizing the variance retained from the different scores (Alnæs et al., 2019; Coombes et al., 2020). Before PCA, all regional MRI ROI measures were residualized for age, sex, scanner and the first 20 ancestry components (see below) using linear models. Similarly, all individual PRS scores were residualized by the first 20 ancestry components. All residuals were then inverse-ranked normally transformed and z-normalized to avoid higher variance variables being overrepresented in the principal components and then submitted to PCA. For each MRI modality, a PCA was performed with the R function *prcomp*, one for cortical thickness, one for surface area, etc. and the first 3 PCs were retained for further analyses. This number of components was chosen arbitrarily to ease interpretability, since the first principal component showed a global measure with all variables loading into it, and the second and third components showed orthogonal regional components. It is important to note that the cumulative variance explained of the first 3 PCs of some modalities was low (<50%, e.g., cortical thickness or RSI-Hind), and therefore the imaging information of some modalities used further explained less than half of the total variance. For that reason, we performed additional tests to ensure the validity of our results. First, we calculated the SNP heritability of all the ROI imaging measures included, as well as the heritability of an extended 20 PC solution per imaging modality as shown in Supplementary materials. Second, we re-assessed the multivariate CCA analysis with 100 PCs of all the MRI data as the imaging input. For the PRS scores, only the first PC was retained because in all cases the variance explained by this component exceeded > 90%, denoted simply as PRS from here on. Component signs were flipped to reflect a positive load on PC1. The individual subject scores for each PC (27 imaging components and 8 PRS) were used for subsequent analyses.

### 2.8 SNP heritability

We used GCTA to quantify the proportion of imaging components and univariate ROI measures variance explained by all the imputed and QCed autosomal SNPs (Yang et al., 2011). Instead of fitting individual SNPs, GCTA fits all common SNPs simultaneously as random effects in a linear mixed model framework. From the QCed and imputed SNP data, pairwise genetic relationships were estimated to generate a Genetic Relationship Matrix (GRM), where each off-diagonal element represents the additive genetic relatedness between pairs of individuals. The GRM from the SNPs was then fitted in a linear mixed model and Restricted Maximum Likelihood was used to estimate the variance explained by all SNPs.

Estimates were tested for significance with a likelihood ratio test, comparing the likelihood of the alternative to that of the null hypothesis. A PCA of the GRM was performed and the eigenvectors of the first 20 PCs (“ancestry PCs”) were used to residualize the imaging and PRS variables. Heritability confidence intervals (CIs) at 95% were estimated using FIESTA (Fast Confidence IntErvals using Stochastic Approximation; (Schweiger et al., 2018, 2017) that uses parametric bootstrapping to calculate accurate heritability CIs based on the eigenvectors and eigenvalues of the GRM, and the same covariates that were used to residualize the MRI data. As a sensitivity step, we repeated the analyses restricting the sample to participants with European genetic ancestry (n=3,841).

### 2.9 Univariate models between imaging components and PRS and CBCL

Linear relationships between imaging components and PRS and CBCL scores were tested with Bayesian models with all standardized brain imaging components as dependent variables and PRS and CBCL scores as independent variables. Bayes factors to quantify the degree of evidence of the alternative (slopes are non-zero) versus the null hypothesis (slopes are 0, intercept only model) were obtained with the *lmFB* R function and were classified in different categories from providing evidence for the null (bf<=0.33), anecdotal evidence suggestive of limited sample size (bf>0.33<=3), to moderate (bf>3<=10), strong (bf>10<=100) and extreme (bf>100) evidence for the alternative hypothesis (45,46). Uncertainty estimates and posterior means were obtained running Bayesian models with *brms* with weakly informative priors (normal priors with mean 0 and standard deviation 0.5), 2000 iterations, 1000 warm-ups and 4 chains. All model parameters were tested for convergence based on the Rhat value (< 1.1).

### 2.10 Canonical Correlation Analysis (CCA)

CCA is a multivariate technique suitable for evaluating associations between two sets of variables. It seeks the latent dimensions that best characterize the shared variance across the original variables. CCA is ideal to decompose the multivariate shared space of complex and high-dimensional brain imaging and non-imaging phenotypes (Miller et al., 2016; Taquet et al., 2021). We used CCA to interrogate the simultaneous associations between brain imaging components on the one hand, and PRS and externalizing and internalizing problems on the other hand. To assess the significance of the canonical modes, we used permutation testing (1000 permutations) by randomly shuffling the rows (participants) of the PRS matrix, thereby breaking the associations between the two variable sets. In each permutation, CCA was performed and the maximum canonical correlation was obtained. The null distribution from this procedure effectively controls for multiple comparisons across all possible modes. The p-values were calculated as the number of canonical correlations from the null distribution that exceeded the observed canonical correlation over the total number of permutations. While the permutation test provides stringent control for family-wise errors, the resulting p-values are ignorant to the generalization performance of the model. To evaluate the generalizability of the modes, we performed a 10-fold cross-validation scheme 100 times as in (Alnæs et al., 2020). For each iteration of the cross-validation loop, each fold was kept out once and 90% of the data was used to calculate the PCA and CCA weights and applied on the kept-out 10% of the sample. An out-of-sample distribution of canonical correlations was calculated by averaging out the correlations across loops. An alternative MRI-PCA solution with 100 PCs derived from all the neuroimaging ROI data (instead of obtaining 3 PCs per imaging modality) was used as input for CCA to test the stability of the CCA modes. Finally, to investigate whether the brain scores from the CCA were useful in predicting other unseen variables from the same individuals, we ran Bayesian linear models with *brms* predicting fluid and crystallized cognition at baseline and follow-up (n observations = 11,051, mean interval=2.01 years). The models included fluid or crystallized uncorrected scores from the NIH toolbox as dependent variables, fixed effects for brain scores, age at baseline, sex, brain scores*time from baseline and random effects for subjects as predictors.

## 3. Results

### 3.1 Imaging components

We used PCA to obtain the first principal components that captured the largest amount of variance of each of the neuroimaging modalities. The first PC of all imaging modalities was a global component with contributions from all ROI measures, and the subsequent PCs showed regionally specific patterns. See Table 1. Full loadings and scree plots are shown in Supplementary Table 2 and Supplementary Figure 1. Figure 1A shows the spatial patterns of the first three components for the cortical measures. The first three components of cortical thickness explained a cumulative 40% of the variance (PC1:32%, PC2:4,5%, PC3:3,5%). PC1 represented a global component, while PC2 and PC3 showed regional components largely following an anterior-posterior and superior-inferior axis, respectively. Similar global to regional decompositions were observed for all imaging modalities. See Supplementary Figures 1-3.

**Figure 1.**
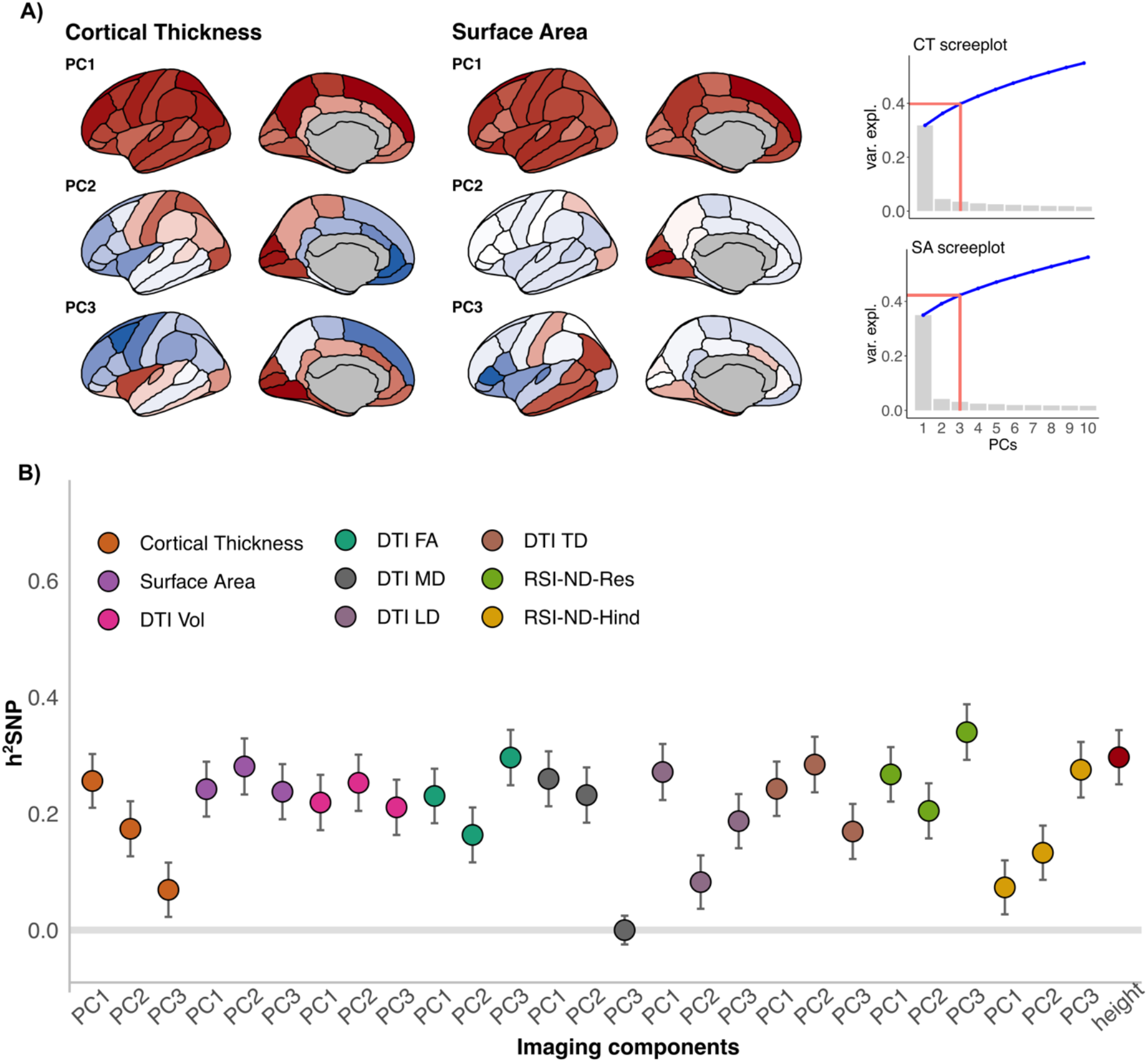
SNP heritability of the first three imaging components of structural neuroimaging data and height. A) Cortical thickness and surface area first three principal components. Brain plots display the loadings of the imaging variables on each of the components. Brain plots were created with *ggseg* (Mowinckel and Vidal-Piñeiro, 2020) and show the left hemisphere for simplicity as loadings were comparable across hemispheres. On the right-hand side, scree plots display the variance (grey bars) and cumulative variance explained (blue line) of the first 10 components. The red line shows the cumulative variance explained by the first 3 components. B) SNP heritability (*Vg/Vp*) point estimates and standard errors (SE) of the imaging components and height. The Y-axis shows the SNP heritability (h^2^SNP). Cortical imaging modalities included cortical thickness and surface area. White matter macro and microstructure components included DTI-Vol (volume of white matter tracts in mm3), DTI-FA (magnitude of directional (spherical) diffusion), DTI-MD (total amount of diffusion), DTI-LD (diffusion parallel to tracts), DTI-TD (diffusion perpendicular to tracts), RSI-Res (normalized directional restricted diffusion, “intracellular”) and RSI-Hind (normalized directional hindered diffusion; “extracellular”).

### 3.2 Heritability of imaging components and ROI measures

We first calculated the SNP heritability of the imaging components and found that most components had moderate heritability estimates (between 0.20 and 0.50). Figure 1B and Supplementary Table S3. 23/27 components remained significant after FDR correction, excluding cortical thickness PC3, DTI-MD PC3, DTI-LD PC2 and RSI-Hind PC1. Similar ranges but moderately higher and more variable estimates were found when restricting the sample to participants of European genetic ancestry (Supplementary Figure 4). For an extended 20 PC solution see Supplementary Figure 5. The top 5 components with the highest point estimates were: RSI-Res PC3 (h^2^SNP=0.34; 95% CI = 0.25-0.44), height (h^2^SNP=0.30; 95% CI = 0.20-0.38), DTI-FA PC3 (h^2^SNP=0.30, 95% CI = 0.21-0.39), DTI-TD PC2 (h^2^SNP=0.28, 95% CI = 0.20-0.37) and surface area PC2 (h^2^SNP=0.28; 95% CI = 0.20-0.37). The heritability of the global components was also moderate. Notably, the global RSI-Hind (normalized directional hindered diffusion of the “extracellular” space”) PC1 had a low heritability and the regional components higher (corticostriatal tracts and fornix). We then calculated the SNP heritability of all univariate ROIs (Figure 2). These results confirmed the imaging component results shown in Figure1B, and showed that RSI-Res (normalized directional restricted diffusion of the “intracellular” space) and DTI-FA of the superior longitudinal fasciculus and the fornix were the modalities and regions with the highest heritability. The average of the non-zero univariate heritability estimates showed that RSI-Res was the modality with the highest heritability and RSI-Hind with the lowest, that all white matter modalities (excluding RSI-Hind) were more heritable than cortical measures and that cortical thickness was marginally less heritable than surface area. Overall, the SNP heritability estimates reported here are comparable albeit lower to some extent to other recent reports in adults (L. T. Elliott et al., 2018; Grasby et al., 2020).

**Figure 2.**
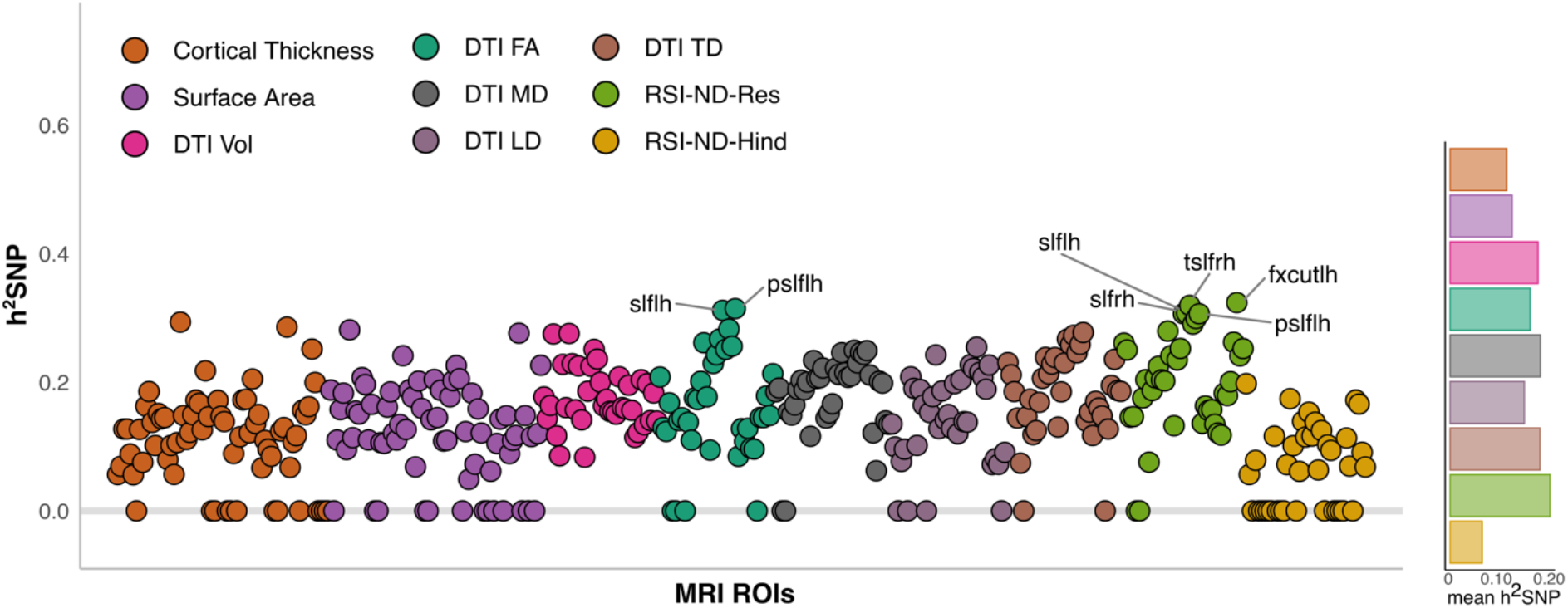
SNP heritability of all bilateral MRI ROI measures. ROIs with the highest heritability point estimates (h2SNP>0.30) are highlighted in the figure. MRI=magnetic resonance imaging; ROIs=regions of interest; h^2^SNP= SNP heritability; slflh= left superior longitudinal fasciculus; slfrh= right superior longitudinal fasciculus; psllh=left posterior part of the superior longitudinal fasciculus; tslfrh = right temporal part of the superior longitudinal fasciculus; fxcutlh=left fornix.

### 3.3. Univariate associations of imaging components, genetic and clinical risk

We then assessed whether imaging components were related to genetic and clinical risk for psychopathology and PRS for educational attainment. These models revealed that the strongest relationships were of imaging components with PRS-Edu and externalizing and internalizing behaviors, further confirmed with Bayes factors suggesting extreme and strong evidence (Figure 3A–B). Higher PRS-Edu were associated with greater surface area PC1, DTI-Vol PC1 and DTI-LD PC1 (Figure 3A), and more externalizing problems with smaller surface area PC1 and DTI-Vol PC1. In contrast, PRS of psychiatric disorders showed overall weaker effects and null to anecdotal evidence. Anecdotal evidence can be indicative of data insensitivity (Dienes, 2014) and suggestive of limited sample size, so it is likely that the ability to detect expected small effects will increase with larger sample sizes. Anecdotal results pointed for example to higher PRS of bipolar disorder associated with greater thickness PC2-PC3, larger surface area PC1 and smaller surface area PC3. Uncertainty intervals are shown in Supplementary Figure 6. When restricting the sample to participants with European genetic ancestry, we found similar relative results with the strongest associations between PRS-Edu and CBCL externalizing problems and global surface area and DTI volumes (see Supplementary Figure 7).

**Figure 3.**
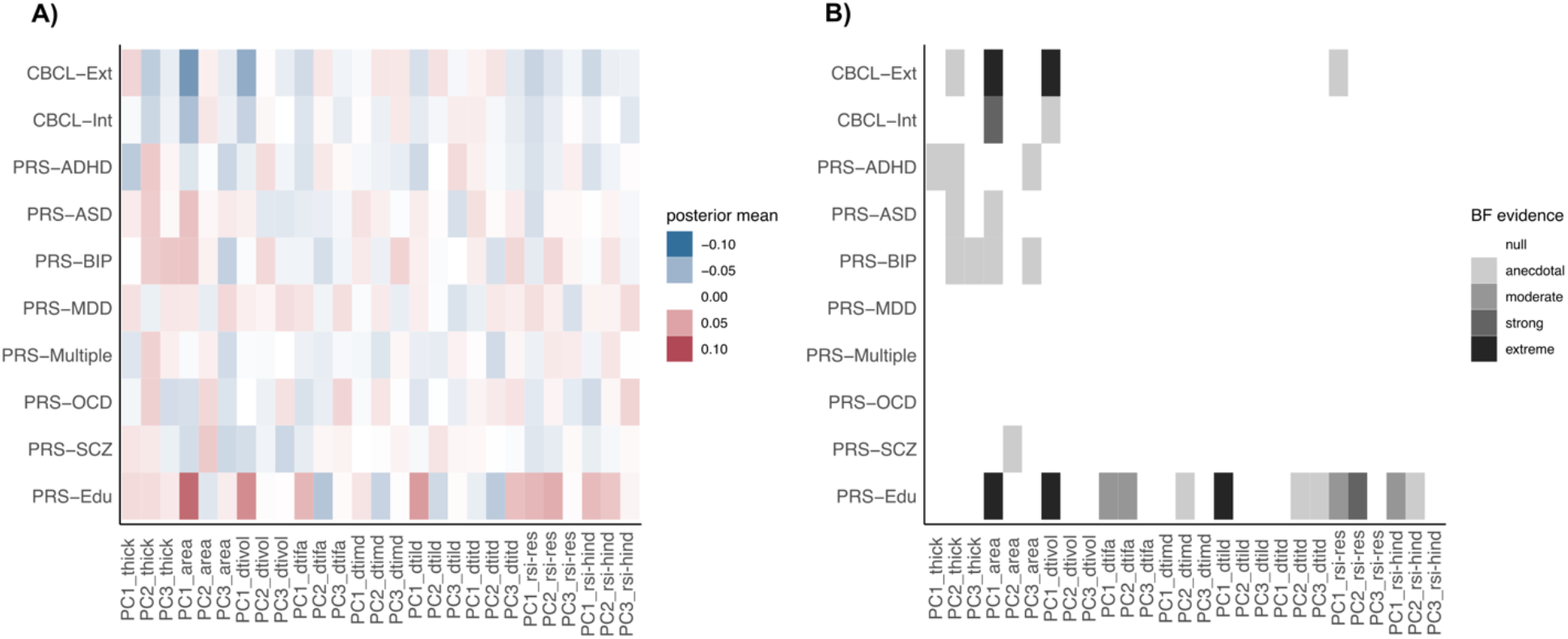
Univariate associations between imaging components and PRS of psychiatric disorders, educational attainment and clinical symptomatology. A) Posterior means of the univariate Bayesian linear regression models between brain imaging (x-axis) and PRS of psychiatric disorders and PRS-Edu and CBCL behavioral scores (y-axis). B) Bayes factors (BF evidence) showed largely null and anecdotal associations between psychiatric PRS and brain imaging components. The strongest evidence showed associations between PRS-Edu, CBCL externalizing problems and global surface area and DTI volumes.

### 3.4 Multivariate patterns of brain imaging, genetic and clinical risk

Finally, we used CCA to evaluate the multivariate patterns of covariation between brain imaging components on the one hand, and PRS of psychiatric disorders and educational attainment and clinical symptomatology on the other. If PRS of psychiatric disorders have distributed effects observed across multiple neuroimaging modalities, then we expect to find significant modes of covariation between PRS and imaging components. We found two significant modes of covariation after permutation testing (Mode1: r=0.153, *p*_perm_<0.001; Mode2: r=0.108, *p*_perm_=0.004). The first mode related higher PRS-Edu and fewer externalizing problems to larger global surface area PC1 and DTI-Vol PC1 (Figure 4). The second orthogonal mode showed that higher PRS of schizophrenia, bipolar disorder and to a lesser extent autism spectrum disorder were associated with higher global cortical thickness PC1, particularly in the occipital cortex, larger surface area of medial occipital regions (PC2), smaller surface area of lateral temporal regions and inferior parietal cortex (PC3) and smaller tract volumes of the fornix, parahippocampal cingulum and forceps major. Although significant, these modes had limited generalizability to unseen data, with canonical correlations in the test sets substantially lower than the empirical correlation observed in the full sample (mean_test_=0.02, mean_traning_=0.13 Figure 4). Brain scores were comparable between white Europeans and other ethnicities (mode1/mode2: posterior mean difference -0.01/0.02; 95% credible interval -0.04-0.05/-0.02-0.07). CCA modes were comparable 1) repeating the analysis without imputing extreme MRI observations (n=7,334; see Supplementary Figure 8) and 2) using 100 principal components from all the MRI data as input to CCA instead of 3 components per modality (correlation brain scores mode1/mode2:0.67/0.54; correlation PRS scores mode1/mode:0.94/0.93).

**Figure 4.**
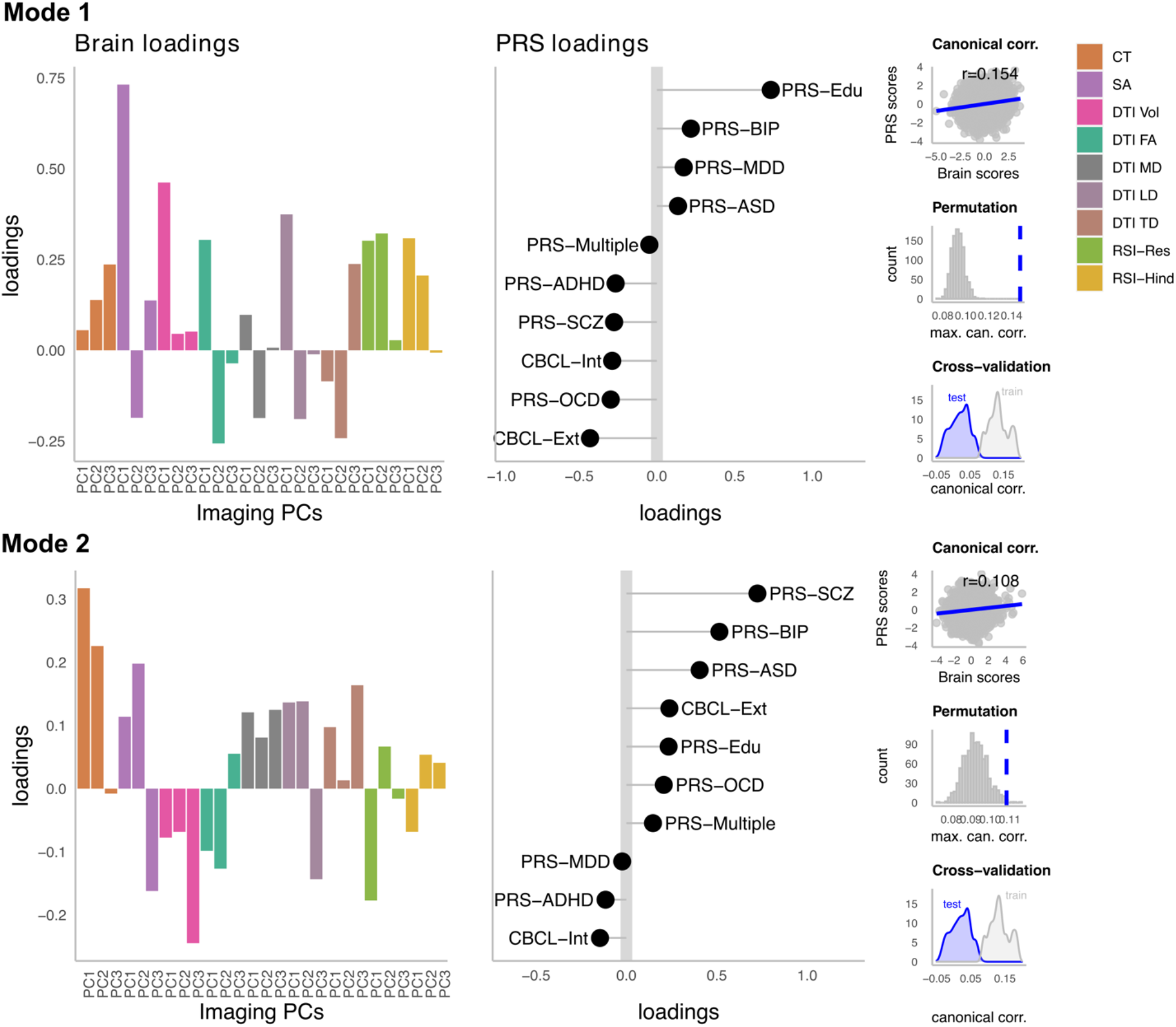
Multivariate modes of covariation between brain imaging and genetic and clinical psychiatric risk. Multivariate canonical modes between brain imaging components and PRS and symptoms were assessed with CCA. The figure shows the loadings (correlation of original variables with the canonical variates) of two significant modes of variation. The first mode (upper row) linked greater global surface area with higher PRS-Edu and fewer externalizing problems. The second mode (bottom row) related greater global cortical thickness and smaller white matter volumes of the fornix and parahippocampal cingulum (PC3) to higher PRS of schizophrenia and bipolar disorder. The right-hand side of both rows shows the canonical correlations, the permuted null distributions and the observed canonical correlations (blue line) and the cross-validation distributions of the average canonical correlations in the test (blue) and training (gray) sets Canonical correlation values in the unseen data (test) were substantially lower than the empirical correlation of the full sample, indicating limited generalizability of the results. The distribution of canonical scores in the training subsets showed comparable canonical correlations to that observed in the full sample. Permutation tests were performed with 1000 permutations to assess the significance of the modes. 10-K cross-validation performed 100 times provided averaged canonical correlation in (training) and out-of-sample (test).

Finally, we tested whether brain scores predicted fluid and crystallized cognitive scores at baseline and their change over a two years interval. We found that Mode 1 brain scores were associated with higher fluid (β=1.04, CI=0.82-1.27) and crystallized scores (β=1.12, CI=0.97-1.28) at baseline, whereas brain scores of Mode 2 two were only marginally associated with lower crystallized scores at baseline (β=-0.19, CI=-0.35--0.04). Brain scores did not predict changes in cognition over time in the subsample examined. See Supplementary Figure 9. These results show that although the multivariate patterns did not generalize well, they were able to capture meaningful variation associated with other unmodeled variables of the same individuals.

## 4. Discussion

Recent evidence from genome-wide and functional genomic studies has put brain development at the foreground of psychiatric disorder’s pathophysiology (Ripke et al., 2020). Here, we combined human neuroimaging with individual SNP data to understand the links between genetic differences and polygenic scores for psychiatric disorders, and the adolescent brain. First, we decomposed each morphological and microstructural imaging modality into their first axes of variation (“imaging components”) and found moderate SNP heritability of most of the components examined. Second, we found small and mostly null associations of imaging components with PRS of psychiatric disorders. The strongest effects revealed higher PRS-Edu with larger surface area PC1 (global). Finally, capitalizing on the interrelated nature of the brain and genetic and clinical risk, we combined brain imaging, genetic and clinical data in a single multivariate model and found two significant modes of covariation after permutation testing. The first mode recapitulated an association between higher PRS-Edu, less externalizing problems and larger surface area PC1. The second orthogonal mode showed that higher PRS of schizophrenia and bipolar disorder were related to higher cortical thickness PC1-PC2, smaller surface area of the lateral temporal and inferior parietal cortex and smaller white matter volumes of the fornix and cingulate tracts. While the stringent cross-validation procedures revealed limited generalizability of the significant multivariate patterns, which could be due to the limited sample size, they agree with current reports showing that the associations between psychiatric polygenic risk and the brain are dominated by small and distributed effects.

We first examined the SNP heritability of imaging components spanning multiple macro and microstructural neuroimaging modalities. We observed modest heritability estimates of most imaging components, conceivably lower than in previous reports with older samples (L. T. Elliott et al., 2018; Zhao et al., 2021). Besides methodological differences, the genetic effects on brain imaging measures are likely to vary across the lifespan, and the heritability of cortical thickness of association areas has been shown to increase throughout adolescence (Lenroot et al., 2009; Schmitt et al., 2014; Teeuw et al., 2019). We also found lower heritability estimates for height than in other adult studies (Yang et al., 2010). Increased heritability with increasing age may be a general phenomenon that applies to many traits (e.g. IQ, (Bouchard, 2013) and could be explained by multiple factors including the decline in environmental variance or gene-environment correlations (Bergen et al., 2007; Schmitt et al., 2014). Imaging components of cortical sensory regions such as the surface area of the pericalcarine, lingual and cuneus ROIs (PC2) were more heritable than associative components, in agreement with prior adolescent and adult studies (Anderson et al., 2021; Schmitt et al., 2019). The regional RSI component reflecting the neurite density of the superior longitudinal fasciculus (RSI-PC3) showed the highest heritability point estimate (h^2^SNP=0.34), further confirmed through univariate analyses (Figure 2). The superior longitudinal fasciculus is an association fiber bundle that connects the occipital, parietal and temporal lobes with the frontal lobes, and matures later than sensory-motor tracts (Bürgel et al., 2006; Westlye et al., 2010). These patterns are in line with recent studies reporting the highest heritability for white matter intracellular volume fraction and diffusivity phenotypes, with the superior longitudinal fasciculus among the highest (Smith et al., 2020; Zhao et al., 2021). In sum, our results highlight the importance for imaging genetics studies to consider the maturational timing of the brain phenotypes examined as well as heterogeneity between imaging modalities and regions.

We then investigated whether PRS of several psychiatric disorders, PRS-Edu and clinical symptomatology were associated with brain imaging components at this age. Using univariate Bayesian linear models, we tested many one-to-one associations between imaging components and PRS and behavioral scores. The strongest effects were found for PRS-Edu, which was related to both global and regional imaging components. Higher PRS-Edu was related to greater surface area PC1 (global) and DTI tract volumes PC1 (global). The relationship between higher PRS-Edu and global surface area in adolescence has been previously shown (Judd et al., 2020), and is consistent with positive genetic correlations between educational attainment and total surface area (Grasby et al., 2020). PRS-Edu was also related to greater regional white matter diffusion of the cortico-striate tracts that connect the frontal and parietal cortices to the striatum (e.g. RSI-Res PC2). Cortico-striatal connectivity appears to greatly develop during adolescence (Palmer et al., 2021), enabling integration between the striatum and the cortex, and possibly subserving cognitive control and value-guided action (Davidow et al., 2018). The relationship of PRS of psychiatric disorders and imaging components were weaker and mostly characterized by null and anecdotal evidence when examined separately for each imaging phenotype. The stronger associations found for PRS-Edu may relate to differences in the original studies that translate into varying predictability. The PRS-Edu currently explains up to 13% of the variance of the phenotype, whereas the PRS-SCZ explains up to 7% (Lee et al., 2018; Ripke et al., 2020).

Our multivariate approach revealed two significant modes of covariation between brain imaging and genetic and clinical risk. The results of this analysis should be interpreted with caution in light of the limited generalizability observed with our cross-validation scheme, and future replication attempts are crucial to determine the implications of these results. Further, small effects may be the norm in larger samples when testing for associations between different complex phenotypes and human traits (Dick et al., 2021; Paulus and Thompson, 2019; Westlye et al., 2019). Even larger sample sizes than the one used here may be needed to be able to detect the expected small effects of psychiatric PRS on the brain (Westlye et al., 2019). The canonical correlations we found were small (r<0.2). Nonetheless, this analysis highlights the added value of using multimodal imaging in a multivariate framework to help understand the associations with the psychiatric risk that would have been missed otherwise. This is more clearly seen in Mode 2. This mode related higher polygenic scores of schizophrenia, bipolar disorder and autism spectrum disorder to higher cortical thickness, smaller surface area in lateral and medial temporal regions (surface area PC3), lower global white matter restricted neurite density (RSI-Res PC1) and smaller white matter volumes in the fornix and cingulum (DTI-Vol PC3). In other words, the PRS for these disorders were not solely related to a single modality or brain area but rather to a combination of morphology and microstructural brain phenotypes. Importantly, these psychiatric PRS were associated with a multimodal brain imaging pattern above and beyond the dimensional clinical symptoms modeled here. In addition to showing higher thickness globally, which is consistent with recent reports in youth samples (Kirschner et al., 2021), it revealed regional shape differences in the lateral and medial temporal cortex, and white matter tracts connecting the hippocampus, a region previously associated with schizophrenia (Alnæs et al., 2019; Harrison, 2004; van der Meer, Dennis et al., 2020). Many biological processes may underlie the observed signals as imaging modalities are sensitive but not specific to single biological processes. It is conceivable that multiple developmental processes (myelination, pruning, morphological changes, etc) are happening in parallel and account for the neuroimaging patterns reported here. Previous animal work points to aberrant pruning in schizophrenia that eventually results in an excess loss of synapses (Sekar, Aswin et al., 2016; Yilmaz et al., 2021). Combining quantitative MRI with diffusion-weighted imaging shows promise to help clarify different processes *in-vivo* in humans (Lazari et al., 2021; Natu et al., 2019). Finally, that the risk for several psychiatric disorders manifests in shared brain correlates supports their transdiagnostic nature (Anttila et al., 2018; M. L. Elliott et al., 2018; Sprooten et al., 2021; Taquet et al., 2021).

Although late childhood and early adolescence represent important developmental periods, prenatal genetic influences continue to be observed in the brain throughout development and aging (Ball et al., 2020; Fjell et al., 2015). Moreover, genes associated with psychiatric disorders show preferential expression during prenatal development (Birnbaum et al., 2014; Jaffe et al., 2018). It is conceivable that at least part of the effects of psychiatric risk on the brain would be observed even earlier in life. Future longitudinal and experimental studies would be beneficial to disentangle developmental and causal relationships. In this study, we included ROI data from multiple modalities, yet high heterogeneity within ROIs has been reported, particularly within tracts and regions with the strongest age effects (Palmer et al., 2021). Multimodal voxel-wise imaging data can be more precise when studying typical and atypical brain development.

Although our main PCA results are consistent with the univariate heritability (Figure 2) and multivariate heritability analyses increasing the number of components (Supplementary Figure 5), our PCA approach only captured gross patterns of variation in the imaging data, and it is plausible that more regionally-specific genetic effects may have been missed. Despite the efforts to minimize sample bias, the ABCD cohort differs from general population estimates (e.g., higher family income, Heeringa and Berglund, 2020) and it is uncertain how this impacts the distributions of the PRS of mental disorders. In addition, more recently developed methods for calculating PRS show higher prediction accuracy than the one employed here and may be more advantageous in psychiatric applications (Ge et al., 2019.; Ni et al., 2021). Finally, heritability is a sample-specific concept, and other sources of variation than genetics e.g. environmental variance and measurement error impact its estimation. Family designs, repeated measures and cross-cultural studies can give a broader view of the factors accompanying brain development.

## 5. Conclusions

To conclude, we found that higher PRS for educational attainment were associated with larger surface area and white matter volumes, confirming previous reports. In contrast, PRS for psychiatric disorders had small and distributed effects on multiple brain imaging measures in late childhood. Our multivariate approach revealed multimodal associations between psychiatric risk and the brain, demonstrating potential pathways for future research and supporting the transdiagnostic nature of psychiatric disorders. Yet, our cross-validation findings act as a reminder of the general knowledge that statistical significance should not be confused with generalization performance, and future studies should test the replicability and generalizability in other populations. Other unmodeled factors such as rare genetic variation and environmental factors may contribute to deepening our understanding of brain development at this age.

## Data Availability

All data produced in the present study are available upon reasonable request to the authors

## Acknowledgments

The European Research Council under the European Union’s Horizon 2020 research and Innovation program (ERC StG, Grant 802998), the Research Council of Norway (223273, 273345, 276082, 298646, 300767), and the South-Eastern Norway Regional Health Authorities (2019101, 2019107, 2020086). Data used in the preparation of this article were obtained from the Adolescent Brain Cognitive Development^SM^ (ABCD) Study (https://abcdstudy.org), held in the NIMH Data Archive (NDA). This is a multisite, longitudinal study designed to recruit more than 10,000 children age 9-10 and follow them over 10 years into early adulthood. The ABCD Study® is supported by the National Institutes of Health and additional federal partners under award numbers U01DA041048, U01DA050989, U01DA051016, U01DA041022, U01DA051018, U01DA051037, U01DA050987, U01DA041174, U01DA041106, U01DA041117, U01DA041028, U01DA041134, U01DA050988, U01DA051039, U01DA041156, U01DA041025, U01DA041120, U01DA051038, U01DA041148, U01DA041093, U01DA041089, U24DA041123, U24DA041147. A full list of supporters is available at https://abcdstudy.org/federal-partners.html. A listing of participating sites and a complete listing of the study investigators can be found at https://abcdstudy.org/consortium_members/. ABCD consortium investigators designed and implemented the study and/or provided data but did not necessarily participate in the analysis or writing of this report. This manuscript reflects the views of the authors and may not reflect the opinions or views of the NIH or ABCD consortium investigators.

## Data and code availability

The ABCD data repository grows and changes over time. The ABCD data used in this report came from NIMH Data Archive Release 3.0 (DOI: http://dx.doi.org/10.15154/1519007). DOIs can be found at https://nda.nih.gov/abcd. The tools used for analyzing the data are publicly available. Genetic data analysis [plink v.1.9: http://pngu.mgh.harvard.edu/purcell/plink/); PRSice-2: https://www.prsice.info/; GCTA v.1.93: https://yanglab.westlake.edu.cn/software/gcta/], statistical analysis and data preprocessing [R v.3.6.3: https://www.r-project.org/]. Scripts for data handling will be made available upon publication at Open Science Framework: https://osf.io/z83mw/?view_only=1fc11f1d2b8449b6ba0d36f589a34b6b

## Author contributions

**S.F.C:** Conceptualization, Methodology, Data curation and analysis, Writing - Original draft, Visualization; **L.T.W:** Conceptualization, Writing - Review and editing, Supervision, Funding acquisition; **D.A**: Conceptualization, Methodology, Data Curation, Writing – Review and editing; **DvdM**: Data Curation, Writing – Review and editing; **A.D**: Writing – Review and editing; **M.H**: Writing – Review and editing; **R.K**: Writing – Review and editing; **I.I.M**: Writing – Review and editing; **L.B.N**: Writing – Review and editing; **M.L.P**: Writing – Review and editing; **I.V**: Writing – Review and editing; **O.A.A**: Writing – Review and editing, Funding acquisition.

## References

Alnæs, D., Kaufmann, T., Marquand, A.F., Smith, S.M., Westlye, L.T., 2020. Patterns of sociocognitive stratification and perinatal risk in the child brain. Proc. Natl. Acad. Sci. 117, 12419–12427. https://doi.org/10.1073/pnas.2001517117

Alnæs, D., Kaufmann, T., van der Meer, D., Córdova-Palomera, A., Rokicki, J., Moberget, T., Bettella, F., Agartz, I., Barch, D.M., Bertolino, A., Brandt, C.L., Cervenka, S., Djurovic, S., Doan, N.T., Eisenacher, S., Fatouros-Bergman, H., Flyckt, L., Di Giorgio, A., Haatveit, B., Jönsson, E.G., Kirsch, P., Lund, M.J., Meyer-Lindenberg, A., Pergola, G., Schwarz, E., Smeland, O.B., Quarto, T., Zink, M., Andreassen, O.A., Westlye, L.T., for the Karolinska Schizophrenia Project Consortium, 2019. Brain Heterogeneity in Schizophrenia and Its Association With Polygenic Risk. JAMA Psychiatry 76, 739. https://doi.org/10.1001/jamapsychiatry.2019.0257

Anderson, K.M., Ge, T., Kong, R., Patrick, L.M., Spreng, R.N., Sabuncu, M.R., Yeo, B.T.T., Holmes, A.J., 2021. Heritability of individualized cortical network topography. Proc. Natl. Acad. Sci. 118, e2016271118. https://doi.org/10.1073/pnas.2016271118

Anttila, V., Bulik-Sullivan, B., Finucane, H.K., Walters, R.K., Bras, J., Duncan, L., Escott-Price, V., Falcone, G.J., Gormley, P., Malik, R., Patsopoulos, N.A., Ripke, S., Wei, Z., Yu, D., Lee, P.H., Turley, P., Grenier-Boley, B., Chouraki, V., Kamatani, Y., Berr, C., Letenneur, L., Hannequin, D., Amouyel, P., Boland, A., Deleuze, J.-F., Duron, E., Vardarajan, B.N., Reitz, C., Goate, A.M., Huentelman, M.J., Kamboh, M.I., Larson, E.B., Rogaeva, E., St George-Hyslop, P., Hakonarson, H., Kukull, W.A., Farrer, L.A., Barnes, L.L., Beach, T.G., Demirci, F.Y., Head, E., Hulette, C.M., Jicha, G.A., Kauwe, J.S.K., Kaye, J.A., Leverenz, J.B., Levey, A.I., Lieberman, A.P., Pankratz, V.S., Poon, W.W., Quinn, J.F., Saykin, A.J., Schneider, L.S., Smith, A.G., Sonnen, J.A., Stern, R.A., Van Deerlin, V.M., Van Eldik, L.J., Harold, D., Russo, G., Rubinsztein, D.C., Bayer, A., Tsolaki, M., Proitsi, P., Fox, N.C., Hampel, H., Owen, M.J., Mead, S., Passmore, P., Morgan, K., Nöthen, M.M., Schott, J.M., Rossor, M., Lupton, M.K., Hoffmann, P., Kornhuber, J., Lawlor, B., McQuillin, A., Al-Chalabi, A., Bis, J.C., Ruiz, A., Boada, M., Seshadri, S., Beiser, A., Rice, K., van der Lee, S.J., De Jager, P.L., Geschwind, D.H., Riemenschneider, M., Riedel-Heller, S., Rotter, J.I., Ransmayr, G., Hyman, B.T., Cruchaga, C., Alegret, M., Winsvold, B., Palta, P., Farh, K.-H., Cuenca-Leon, E., Furlotte, N., Kurth, T., Ligthart, L., Terwindt, G.M., Freilinger, T., Ran, C., Gordon, S.D., Borck, G., Adams, H.H.H., Lehtimäki, T., Wedenoja, J., Buring, J.E., Schürks, M., Hrafnsdottir, M., Hottenga, J.-J., Penninx, B., Artto, V., Kaunisto, M., Vepsäläinen, S., Martin, N.G., Montgomery, G.W., Kurki, M.I., Hämäläinen, E., Huang, H., Huang, J., Sandor, C., Webber, C., Muller-Myhsok, B., Schreiber, S., Salomaa, V., Loehrer, E., Göbel, H., Macaya, A., Pozo-Rosich, P., Hansen, T., Werge, T., Kaprio, J., Metspalu, A., Kubisch, C., Ferrari, M.D., Belin, A.C., van den Maagdenberg, A.M.J.M., Zwart, J.-A., Boomsma, D., Eriksson, N., Olesen, J., Chasman, D.I., Nyholt, D.R., Anney, R., Avbersek, A., Baum, L., Berkovic, S., Bradfield, J., Buono, R.J., Catarino, C.B., Cossette, P., De Jonghe, P., Depondt, C., Dlugos, D., Ferraro, T.N., French, J., Hjalgrim, H., Jamnadas-Khoda, J., Kälviäinen, R., Kunz, W.S., Lerche, H., Leu, C., Lindhout, D., Lo, W., Lowenstein, D., McCormack, M., Møller, R.S., Molloy, A., Ng, P.-W., Oliver, K., Privitera, M., Radtke, R., Ruppert, A.-K., Sander, T., Schachter, S., Schankin, C., Scheffer, I., Schoch, S., Sisodiya, S.M., Smith, P., Sperling, M., Striano, P., Surges, R., Thomas, G.N., Visscher, F., Whelan, C.D., Zara, F., Heinzen, E.L., Marson, A., Becker, F., Stroink, H., Zimprich, F., Gasser, T., Gibbs, R., Heutink, P., Martinez, M., Morris, H.R., Sharma, M., Ryten, M., Mok, K.Y., Pulit, S., Bevan, S., Holliday, E., Attia, J., Battey, T., Boncoraglio, G., Thijs, V., Chen, W.-M., Mitchell, B., Rothwell, P., Sharma, P., Sudlow, C., Vicente, A., Markus, H., Kourkoulis, C., Pera, J., Raffeld, M., Silliman, S., Boraska Perica, V., Thornton, L.M., Huckins, L.M., William Rayner, N., Lewis, C.M., Gratacos, M., Rybakowski, F., Keski-Rahkonen, A., Raevuori, A., Hudson, J.I., Reichborn-Kjennerud, T., Monteleone, P., Karwautz, A., Mannik, K., Baker, J.H., O’Toole, J.K., Trace, S.E., Davis, O.S.P., Helder, S.G., Ehrlich, S., Herpertz-Dahlmann, B., Danner, U.N., van Elburg, A.A., Clementi, M., Forzan, M., Docampo, E., Lissowska, J., Hauser, J., Tortorella, A., Maj, M., Gonidakis, F., Tziouvas, K., Papezova, H., Yilmaz, Z., Wagner, G., Cohen-Woods, S., Herms, S., Julià, A., Rabionet, R., Dick, D.M., Ripatti, S., Andreassen, O.A., Espeseth, T., Lundervold, A.J., Steen, V.M., Pinto, D., Scherer, S.W., Aschauer, H., Schosser, A., Alfredsson, L., Padyukov, L., Halmi, K.A., Mitchell, J., Strober, M., Bergen, A.W., Kaye, W., Szatkiewicz, J.P., Cormand, B., Ramos-Quiroga, J.A., Sánchez-Mora, C., Ribasés, M., Casas, M., Hervas, A., Arranz, M.J., Haavik, J., Zayats, T., Johansson, S., Williams, N., Elia, J., Dempfle, A., Rothenberger, A., Kuntsi, J., Oades, R.D., Banaschewski, T., Franke, B., Buitelaar, J.K., Arias Vasquez, A., Doyle, A.E., Reif, A., Lesch, K.-P., Freitag, C., Rivero, O., Palmason, H., Romanos, M., Langley, K., Rietschel, M., Witt, S.H., Dalsgaard, S., Børglum, A.D., Waldman, I., Wilmot, B., Molly, N., Bau, C.H.D., Crosbie, J., Schachar, R., Loo, S.K., McGough, J.J., Grevet, E.H., Medland, S.E., Robinson, E., Weiss, L.A., Bacchelli, E., Bailey, A., Bal, V., Battaglia, A., Betancur, C., Bolton, P., Cantor, R., Celestino-Soper, P., Dawson, G., De Rubeis, S., Duque, F., Green, A., Klauck, S.M., Leboyer, M., Levitt, P., Maestrini, E., Mane, S., De-Luca, D.M.-, Parr, J., Regan, R., Reichenberg, A., Sandin, S., Vorstman, J., Wassink, T., Wijsman, E., Cook, E., Santangelo, S., Delorme, R., Rogé, B., Magalhaes, T., Arking, D., Schulze, T.G., Thompson, R.C., Strohmaier, J., Matthews, K., Melle, I., Morris, D., Blackwood, D., McIntosh, A., Bergen, S.E., Schalling, M., Jamain, S., Maaser, A., Fischer, S.B., Reinbold, C.S., Fullerton, J.M., Grigoroiu-Serbanescu, M., Guzman-Parra, J., Mayoral, F., Schofield, P.R., Cichon, S., Mühleisen, T.W., Degenhardt, F., Schumacher, J., Bauer, M., Mitchell, P.B., Gershon, E.S., Rice, J., Potash, J.B., Zandi, P.P., Craddock, N., Ferrier, I.N., Alda, M., Rouleau, G.A., Turecki, G., Ophoff, R., Pato, C., Anjorin, A., Stahl, E., Leber, M., Czerski, P.M., Edenberg, H.J., Cruceanu, C., Jones, I.R., Posthuma, D., Andlauer, T.F.M., Forstner, A.J., Streit, F., Baune, B.T., Air, T., Sinnamon, G., Wray, N.R., MacIntyre, D.J., Porteous, D., Homuth, G., Rivera, M., Grove, J., Middeldorp, C.M., Hickie, I., Pergadia, M., Mehta, D., Smit, J.H., Jansen, R., de Geus, E., Dunn, E., Li, Q.S., Nauck, M., Schoevers, R.A., Beekman, A.T., Knowles, J.A., Viktorin, A., Arnold, P., Barr, C.L., Bedoya-Berrio, G., Bienvenu, O.J., Brentani, H., Burton, C., Camarena, B., Cappi, C., Cath, D., Cavallini, M., Cusi, D., Darrow, S., Denys, D., Derks, E.M., Dietrich, A., Fernandez, T., Figee, M., Freimer, N., Gerber, G., Grados, M., Greenberg, E., Hanna, G.L., Hartmann, A., Hirschtritt, M.E., Hoekstra, P.J., Huang, A., Huyser, C., Illmann, C., Jenike, M., Kuperman, S., Leventhal, B., Lochner, C., Lyon, G.J., Macciardi, F., Madruga-Garrido, M., Malaty, I.A., Maras, A., McGrath, L., Miguel, E.C., Mir, P., Nestadt, G., Nicolini, H., Okun, M.S., Pakstis, A., Paschou, P., Piacentini, J., Pittenger, C., Plessen, K., Ramensky, V., Ramos, E.M., Reus, V., Richter, M.A., Riddle, M.A., Robertson, M.M., Roessner, V., Rosário, M., Samuels, J.F., Sandor, P., Stein, D.J., Tsetsos, F., Van Nieuwerburgh, F., Weatherall, S., Wendland, J.R., Wolanczyk, T., Worbe, Y., Zai, G., Goes, F.S., McLaughlin, N., Nestadt, P.S., Grabe, H.-J., Depienne, C., Konkashbaev, A., Lanzagorta, N., Valencia-Duarte, A., Bramon, E., Buccola, N., Cahn, W., Cairns, M., Chong, S.A., Cohen, D., Crespo-Facorro, B., Crowley, J., Davidson, M., DeLisi, L., Dinan, T., Donohoe, G., Drapeau, E., Duan, J., Haan, L., Hougaard, D., Karachanak-Yankova, S., Khrunin, A., Klovins, J., Kučinskas, V., Lee Chee Keong, J., Limborska, S., Loughland, C., Lönnqvist, J., Maher, B., Mattheisen, M., McDonald, C., Murphy, K.C., Murray, R., Nenadic, I., van Os, J., Pantelis, C., Pato, M., Petryshen, T., Quested, D., Roussos, P., Sanders, A.R., Schall, U., Schwab, S.G., Sim, K., So, H.-C., Stögmann, E., Subramaniam, M., Toncheva, D., Waddington, J., Walters, J., Weiser, M., Cheng, W., Cloninger, R., Curtis, D., Gejman, P.V., Henskens, F., Mattingsdal, M., Oh, S.-Y., Scott, R., Webb, B., Breen, G., Churchhouse, C., Bulik, C.M., Daly, M., Dichgans, M., Faraone, S.V., Guerreiro, R., Holmans, P., Kendler, K.S., Koeleman, B., Mathews, C.A., Price, A., Scharf, J., Sklar, P., Williams, J., Wood, N.W., Cotsapas, C., Palotie, A., Smoller, J.W., Sullivan, P., Rosand, J., Corvin, A., Neale, B.M., 2018. Analysis of shared heritability in common disorders of the brain. Science 360, eaap8757. https://doi.org/10.1126/science.aap8757

Ball, G., Seidlitz, J., O’Muircheartaigh, J., Dimitrova, R., Fenchel, D., Makropoulos, A., Christiaens, D., Schuh, A., Passerat-Palmbach, J., Hutter, J., Cordero-Grande, L., Hughes, E., Price, A., Hajnal, J.V., Rueckert, D., Robinson, E.C., Edwards, A.D., 2020. Cortical morphology at birth reflects spatiotemporal patterns of gene expression in the fetal human brain. PLOS Biol. 18, e3000976. https://doi.org/10.1371/journal.pbio.3000976

Baurley, J.W., Edlund, C.K., Pardamean, C.I., Conti, D.V., Bergen, A.W., 2016. Smokescreen: a targeted genotyping array for addiction research. BMC Genomics 17. https://doi.org/10.1186/s12864-016-2495-7

Bergen, S.E., Gardner, C.O., Kendler, K.S., 2007. Age-Related Changes in Heritability of Behavioral Phenotypes Over Adolescence and Young Adulthood: A Meta-Analysis. Twin Res. Hum. Genet. 10, 423–433. https://doi.org/10.1375/twin.10.3.423

Birnbaum, R., Jaffe, A.E., Hyde, T.M., Kleinman, J.E., Weinberger, D.R., 2014. Prenatal Expression Patterns of Genes Associated With Neuropsychiatric Disorders. Am. J. Psychiatry 171, 758–767. https://doi.org/10.1176/appi.ajp.2014.13111452

Birnbaum, R., Weinberger, D.R., 2017. Genetic insights into the neurodevelopmental origins of schizophrenia. Nat. Rev. Neurosci. 18, 727–740. https://doi.org/10.1038/nrn.2017.125

Bouchard, T.J., 2013. The Wilson Effect: The Increase in Heritability of IQ With Age. Twin Res. Hum. Genet. 16, 923–930. https://doi.org/10.1017/thg.2013.54

Bürgel, U., Amunts, K., Hoemke, L., Mohlberg, H., Gilsbach, J.M., Zilles, K., 2006. White matter fiber tracts of the human brain: Three-dimensional mapping at microscopic resolution, topography and intersubject variability. NeuroImage 29, 1092–1105. https://doi.org/10.1016/j.neuroimage.2005.08.040

Casey, B.J., Cannonier, T., Conley, M.I., Cohen, A.O., Barch, D.M., Heitzeg, M.M., Soules, M.E., Teslovich, T., Dellarco, D.V., Garavan, H., Orr, C.A., Wager, T.D., Banich, M.T., Speer, N.K., Sutherland, M.T., Riedel, M.C., Dick, A.S., Bjork, J.M., Thomas, K.M., Chaarani, B., Mejia, M.H., Hagler, D.J., Daniela Cornejo, M., Sicat, C.S., Harms, M.P., Dosenbach, N.U.F., Rosenberg, M., Earl, E., Bartsch, H., Watts, R., Polimeni, J.R., Kuperman, J.M., Fair, D.A., Dale, A.M., ABCD Imaging Acquisition Workgroup, 2018. The Adolescent Brain Cognitive Development (ABCD) study: Imaging acquisition across 21 sites. Dev. Cogn. Neurosci. 32, 43–54. https://doi.org/10.1016/j.dcn.2018.03.001

Choi, S.W., O’Reilly, P.F., 2019. PRSice-2: Polygenic Risk Score software for biobank-scale data. GigaScience 8, giz082. https://doi.org/10.1093/gigascience/giz082

Colby, J.B., Van Horn, J.D., Sowell, E.R., 2011. Quantitative in vivo evidence for broad regional gradients in the timing of white matter maturation during adolescence. NeuroImage 54, 25–31. https://doi.org/10.1016/j.neuroimage.2010.08.014

Coombes, B.J., Ploner, A., Bergen, S.E., Biernacka, J.M., 2020. A principal component approach to improve association testing with polygenic risk scores. Genet. Epidemiol. 44, 676–686. https://doi.org/10.1002/gepi.22339

Cuthbert, B.N., 2014. The RDoC framework: facilitating transition from ICD/DSM to dimensional approaches that integrate neuroscience and psychopathology: Forum - The Research Domain Criteria Project. World Psychiatry 13, 28–35. https://doi.org/10.1002/wps.20087

Dahl, R.E., Allen, N.B., Wilbrecht, L., Suleiman, A.B., 2018. Importance of investing in adolescence from a developmental science perspective. Nature 554, 441–450. https://doi.org/10.1038/nature25770

Davidow, J.Y., Insel, C., Somerville, L.H., 2018. Adolescent Development of Value-Guided Goal Pursuit. Trends Cogn. Sci. 22, 725–736. https://doi.org/10.1016/j.tics.2018.05.003

Demontis, D., Walters, R.K., Martin, J., Mattheisen, M., Als, T.D., Agerbo, E., Baldursson, G., Belliveau, R., Bybjerg-Grauholm, J., Bækvad-Hansen, M., Cerrato, F., Chambert, K., Churchhouse, C., Dumont, A., Eriksson, N., Gandal, M., Goldstein, J.I., Grasby, K.L., Grove, J., Gudmundsson, O.O., Hansen, C.S., Hauberg, M.E., Hollegaard, M.V., Howrigan, D.P., Huang, H., Maller, J.B., Martin, A.R., Martin, N.G., Moran, J., Pallesen, J., Palmer, D.S., Pedersen, C.B., Pedersen, M.G., Poterba, T., Poulsen, J.B., Ripke, S., Robinson, E.B., Satterstrom, F.K., Stefansson, H., Stevens, C., Turley, P., Walters, G.B., Won, H., Wright, M.J., Andreassen, O.A., Asherson, P., Burton, C.L., Boomsma, D.I., Cormand, B., Dalsgaard, S., Franke, B., Gelernter, J., Geschwind, D., Hakonarson, H., Haavik, J., Kranzler, H.R., Kuntsi, J., Langley, K., Lesch, K.-P., Middeldorp, C., Reif, A., Rohde, L.A., Roussos, P., Schachar, R., Sklar, P., Sonuga-Barke, E.J.S., Sullivan, P.F., Thapar, A., Tung, J.Y., Waldman, I.D., Medland, S.E., Stefansson, K., Nordentoft, M., Hougaard, D.M., Werge, T., Mors, O., Mortensen, P.B., Daly, M.J., Faraone, S.V., Børglum, A.D., Neale, B.M., 2019. Discovery of the first genome-wide significant risk loci for attention deficit/hyperactivity disorder. Nat. Genet. 51, 63–75. https://doi.org/10.1038/s41588-018-0269-7

Desikan, R.S., Ségonne, F., Fischl, B., Quinn, B.T., Dickerson, B.C., Blacker, D., Buckner, R.L., Dale, A.M., Maguire, R.P., Hyman, B.T., Albert, M.S., Killiany, R.J., 2006. An automated labeling system for subdividing the human cerebral cortex on MRI scans into gyral based regions of interest. NeuroImage 31, 968–980. https://doi.org/10.1016/j.neuroimage.2006.01.021

Dick, A.S., Lopez, D.A., Watts, A.L., Heeringa, S., Reuter, C., Bartsch, H., Fan, C.C., Kennedy, D.N., Palmer, C., Marshall, A., Haist, F., Hawes, S., Nichols, T.E., Barch, D.M., Jernigan, T.L., Garavan, H., Grant, S., Pariyadath, V., Hoffman, E., Neale, M., Stuart, E.A., Paulus, M.P., Sher, K.J., Thompson, W.K., 2021. Meaningful associations in the adolescent brain cognitive development study. NeuroImage 239, 118262. https://doi.org/10.1016/j.neuroimage.2021.118262

Dienes, Z., 2014. Using Bayes to get the most out of non-significant results. Front. Psychol. 5. https://doi.org/10.3389/fpsyg.2014.00781

Du, J., Buckner, R.L., 2021. Precision estimates of macroscale network organization in the human and their relation to anatomical connectivity in the marmoset monkey. Curr. Opin. Behav. Sci. 40, 144–152. https://doi.org/10.1016/j.cobeha.2021.04.010

Elliott, L.T., Sharp, K., Alfaro-Almagro, F., Shi, S., Miller, K.L., Douaud, G., Marchini, J., Smith, S.M., 2018. Genome-wide association studies of brain imaging phenotypes in UK Biobank. Nature 562, 210–216. https://doi.org/10.1038/s41586-018-0571-7

Elliott, M.L., Romer, A., Knodt, A.R., Hariri, A.R., 2018. A Connectome-wide Functional Signature of Transdiagnostic Risk for Mental Illness. Biol. Psychiatry 84, 452–459. https://doi.org/10.1016/j.biopsych.2018.03.012

Fjell, A.M., Grydeland, H., Krogsrud, S.K., Amlien, I., Rohani, D.A., Ferschmann, L., Storsve, A.B., Tamnes, C.K., Sala-Llonch, R., Due-Tønnessen, P., Bjørnerud, A., Sølsnes, A.E., Håberg, A.K., Skranes, J., Bartsch, H., Chen, C.-H., Thompson, W.K., Panizzon, M.S., Kremen, W.S., Dale, A.M., Walhovd, K.B., 2015. Development and aging of cortical thickness correspond to genetic organization patterns. Proc. Natl. Acad. Sci. 112, 15462–15467. https://doi.org/10.1073/pnas.1508831112

Ge, T., Chen, C.-Y., Ni, Y., Anne Feng, Y.-C., Smoller, J.W., n.d. Polygenic prediction via Bayesian regression and continuous shrinkage priors. Nat. Commun. 10.1, 1–10.

Grasby, K.L., Jahanshad, N., Painter, J.N., Colodro-Conde, L., Bralten, J., Hibar, D.P., Lind, P.A., Pizzagalli, F., Ching, C.R.K., McMahon, M.A.B., Shatokhina, N., Zsembik, L.C.P., Thomopoulos, S.I., Zhu, A.H., Strike, L.T., Agartz, I., Alhusaini, S., Almeida, M.A.A., Alnæs, D., Amlien, I.K., Andersson, M., Ard, T., Armstrong, N.J., Ashley-Koch, A., Atkins, J.R., Bernard, M., Brouwer, R.M., Buimer, E.E.L., Bülow, R., Bürger, C., Cannon, D.M., Chakravarty, M., Chen, Q., Cheung, J.W., Couvy-Duchesne, B., Dale, A.M., Dalvie, S., Araujo, T.K. de, Zubicaray, G.I. de, Zwarte, S.M.C. de, Braber, A. den, Doan, N.T., Dohm, K., Ehrlich, S., Engelbrecht, H.-R., Erk, S., Fan, C.C., Fedko, I.O., Foley, S.F., Ford, J.M., Fukunaga, M., Garrett, M.E., Ge, T., Giddaluru, S., Goldman, A.L., Green, M.J., Groenewold, N.A., Grotegerd, D., Gurholt, T.P., Gutman, B.A., Hansell, N.K., Harris, M.A., Harrison, M.B., Haswell, C.C., Hauser, M., Herms, S., Heslenfeld, D.J., Ho, N.F., Hoehn, D., Hoffmann, P., Holleran, L., Hoogman, M., Hottenga, J.-J., Ikeda, M., Janowitz, D., Jansen, I.E., Jia, T., Jockwitz, C., Kanai, R., Karama, S., Kasperaviciute, D., Kaufmann, T., Kelly, S., Kikuchi, M., Klein, M., Knapp, M., Knodt, A.R., Krämer, B., Lam, M., Lancaster, T.M., Lee, P.H., Lett, T.A., Lewis, L.B., Lopes-Cendes, I., Luciano, M., Macciardi, F., Marquand, A.F., Mathias, S.R., Melzer, T.R., Milaneschi, Y., Mirza-Schreiber, N., Moreira, J.C.V., Mühleisen, T.W., Müller-Myhsok, B., Najt, P., Nakahara, S., Nho, K., Loohuis, L.M.O., Orfanos, D.P., Pearson, J.F., Pitcher, T.L., Pütz, B., Quidé, Y., Ragothaman, A., Rashid, F.M., Reay, W.R., Redlich, R., Reinbold, C.S., Repple, J., Richard, G., Riedel, B.C., Risacher, S.L., Rocha, C.S., Mota, N.R., Salminen, L., Saremi, A., Saykin, A.J., Schlag, F., Schmaal, L., Schofield, P.R., Secolin, R., Shapland, C.Y., Shen, L., Shin, J., Shumskaya, E., Sønderby, I.E., Sprooten, E., Tansey, K.E., Teumer, A., Thalamuthu, A., Tordesillas-Gutiérrez, D., Turner, J.A., Uhlmann, A., Vallerga, C.L. Meer, D. van der, Donkelaar, M.M.J. van, Eijk, L. van, Erp, T.G.M. van, Haren, N.E.M. van, Rooij, D. van, Tol, M.-J. van, Veldink, J.H., Verhoef, E., Walton, E., Wang, M., Wang, Y., Wardlaw, J.M., Wen, W., Westlye, L.T., Whelan, C.D., Witt, S.H., Wittfeld, K., Wolf, C., Wolfers, T., Wu, J.Q., Yasuda, C.L., Zaremba, D., Zhang, Z., Zwiers, M.P., Artiges, E., Assareh, A.A., Ayesa-Arriola, R., Belger, A., Brandt, C.L., Brown, G.G., Cichon, S., Curran, J.E., Davies, G.E., Degenhardt, F., Dennis, M.F., Dietsche, B., Djurovic, S., Doherty, C.P., Espiritu, R., Garijo, D., Gil, Y., Gowland, P.A., Green, R.C., Häusler, A.N., Heindel, W., Ho, B.-C., Hoffmann, W.U., Holsboer, F., Homuth, G., Hosten, N., Jack, C.R., Jang, M., Jansen, A., Kimbrel, N.A., Kolskår, K., Koops, S., Krug, A., Lim, K.O., Luykx, J.J., Mathalon, D.H., Mather, K.A., Mattay, V.S., Matthews, S., Son, J.M.V., McEwen, S.C., Melle, I., Morris, D.W., Mueller, B.A., Nauck, M., Nordvik, J.E., Nöthen, M.M., O’Leary, D.S., Opel, N., Martinot, M.-L.P., Pike, G.B., Preda, A., Quinlan, E.B., Rasser, P.E., Ratnakar, V., Reppermund, S., Steen, V.M., Tooney, P.A., Torres, F.R., Veltman, D.J., Voyvodic, J.T., Whelan, R., White, T., Yamamori, H., Adams, H.H.H., Bis, J.C., Debette, S., Decarli, C., Fornage, M., Gudnason, V., Hofer, E., Ikram, M.A., Launer, L., Longstreth, W.T., Lopez, O.L., Mazoyer, B., Mosley, T.H., Roshchupkin, G.V., Satizabal, C.L., Schmidt, R., Seshadri, S., Yang, Q., Initiative¶, A.D.N., Consortium¶, C., Consortium¶, E., Consortium¶, I., Consortium¶, S.Y.S., Initiative¶, P.P.M., Alvim, M.K.M., Ames, D., Anderson, T.J., Andreassen, O.A., Arias-Vasquez, A., Bastin, M.E., Baune, B.T., Beckham, J.C., Blangero, J., Boomsma, D.I., Brodaty, H., Brunner, H.G., Buckner, R.L., Buitelaar, J.K., Bustillo, J.R., Cahn, W., Cairns, M.J., Calhoun, V., Carr, V.J., Caseras, X., Caspers, S., Cavalleri, G.L., Cendes, F., Corvin, A., Crespo-Facorro, B., Dalrymple-Alford, J.C., Dannlowski, U., Geus, E.J.C. de, Deary, I.J., Delanty, N., Depondt, C., Desrivières, S., Donohoe, G., Espeseth, T., Fernández, G., Fisher, S.E., Flor, H., Forstner, A.J., Francks, C., Franke, B., Glahn, D.C., Gollub, R.L., Grabe, H.J., Gruber, O., Håberg, A.K., Hariri, A.R., Hartman, C.A., Hashimoto, R., Heinz, A., Henskens, F.A., Hillegers, M.H.J., Hoekstra, P.J., Holmes, A.J., Hong, L.E., Hopkins, W.D., Pol, H.E.H., Jernigan, T.L., Jönsson, E.G., Kahn, R.S., Kennedy, M.A., Kircher, T.T.J., Kochunov, P., Kwok, J.B.J., Hellard, S.L., Loughland, C.M., Martin, N.G., Martinot, J.-L., McDonald, C., McMahon, K.L., Meyer-Lindenberg, A., Michie, P.T., Morey, R.A., Mowry, B., Nyberg, L., Oosterlaan, J., Ophoff, R.A., Pantelis, C., Paus, T., Pausova, Z., Penninx, B.W.J.H., Polderman, T.J.C., Posthuma, D., Rietschel, M., Roffman, J.L., Rowland, L.M., Sachdev, P.S., Sämann, P.G., Schall, U., Schumann, G., Scott, R.J., Sim, K., Sisodiya, S.M., Smoller, J.W., Sommer, I.E., Pourcain, B.S., Stein, D.J., Toga, A.W., Trollor, J.N., Wee, N.J.A.V. der, Ent, D. van ‘t, Völzke, H., Walter, H., Weber, B., Weinberger, D.R., Wright, M.J., Zhou, J., Stein, J.L., Thompson, P.M., Medland, S.E., Group, E.N.G. through M.-A.C. (ENIGMA)—Genetics working, 2020. The genetic architecture of the human cerebral cortex. Science 367. https://doi.org/10.1126/science.aay6690

Hagler, D.J., Ahmadi, M.E., Kuperman, J., Holland, D., McDonald, C.R., Halgren, E., Dale, A.M., 2009. Automated white-matter tractography using a probabilistic diffusion tensor atlas: Application to temporal lobe epilepsy. Hum. Brain Mapp. 30, 1535–1547. https://doi.org/10.1002/hbm.20619

Hagler, D.J., Hatton, SeanN., Cornejo, M.D., Makowski, C., Fair, D.A., Dick, A.S., Sutherland, M.T., Casey, B.J., Barch, D.M., Harms, M.P., Watts, R., Bjork, J.M., Garavan, H.P., Hilmer, L., Pung, C.J., Sicat, C.S., Kuperman, J., Bartsch, H., Xue, F., Heitzeg, M.M., Laird, A.R., Trinh, T.T., Gonzalez, R., Tapert, S.F., Riedel, M.C., Squeglia, L.M., Hyde, L.W., Rosenberg, M.D., Earl, E.A., Howlett, K.D., Baker, F.C., Soules, M., Diaz, J., de Leon, O.R., Thompson, W.K., Neale, M.C., Herting, M., Sowell, E.R., Alvarez, R.P., Hawes, S.W., Sanchez, M., Bodurka, J., Breslin, F.J., Morris, A.S., Paulus, M.P., Simmons, W.K., Polimeni, J.R., van der Kouwe, A., Nencka, A.S., Gray, K.M., Pierpaoli, C., Matochik, J.A., Noronha, A., Aklin, W.M., Conway, K., Glantz, M., Hoffman, E., Little, R., Lopez, M., Pariyadath, V., Weiss, S.RB., Wolff-Hughes, D.L., DelCarmen-Wiggins, R., Feldstein Ewing, S.W., Miranda-Dominguez, O., Nagel, B.J., Perrone, A.J., Sturgeon, D.T., Goldstone, A., Pfefferbaum, A., Pohl, K.M., Prouty, D., Uban, K., Bookheimer, S.Y., Dapretto, M., Galvan, A., Bagot, K., Giedd, J., Infante, M.A., Jacobus, J., Patrick, K., Shilling, P.D., Desikan, R., Li, Y., Sugrue, L., Banich, M.T., Friedman, N., Hewitt, J.K., Hopfer, C., Sakai, J., Tanabe, J., Cottler, L.B., Nixon, S.J., Chang, L., Cloak, C., Ernst, T., Reeves, G., Kennedy, D.N., Heeringa, S., Peltier, S., Schulenberg, J., Sripada, C., Zucker, R.A., Iacono, W.G., Luciana, M., Calabro, F.J., Clark, D.B., Lewis, D.A., Luna, B., Schirda, C., Brima, T., Foxe, J.J., Freedman, E.G., Mruzek, D.W., Mason, M.J., Huber, R., McGlade, E., Prescot, A., Renshaw, P.F., Yurgelun-Todd, D.A., Allgaier, N.A., Dumas, J.A., Ivanova, M., Potter, A., Florsheim, P., Larson, C., Lisdahl, K., Charness, M.E., Fuemmeler, B., Hettema, J.M., Maes, H.H., Steinberg, J., Anokhin, A.P., Glaser, P., Heath, A.C., Madden, P.A., Baskin-Sommers, A., Constable, R.T., Grant, S.J., Dowling, G.J., Brown, S.A., Jernigan, T.L., Dale, A.M., 2019. Image processing and analysis methods for the Adolescent Brain Cognitive Development Study. NeuroImage 202, 116091. https://doi.org/10.1016/j.neuroimage.2019.116091

Harrison, PaulJ., 2004. The hippocampus in schizophrenia: a review of the neuropathological evidence and its pathophysiological implications. Psychopharmacology (Berl.) 174. https://doi.org/10.1007/s00213-003-1761-y

Heeringa, S.G., Berglund, P.A., 2020. A Guide for Population-based Analysis of the Adolescent Brain Cognitive Development (ABCD) Study Baseline Data (preprint). Neuroscience. https://doi.org/10.1101/2020.02.10.942011

Huntenburg, J.M., Bazin, P.-L., Margulies, D.S., 2018. Large-Scale Gradients in Human Cortical Organization. Trends Cogn. Sci. 22, 21–31. https://doi.org/10.1016/j.tics.2017.11.002

Insel, T.R., 2010. Rethinking schizophrenia. Nature 468, 187–193. https://doi.org/10.1038/nature09552

International Obsessive Compulsive Disorder Foundation Genetics Collaborative (IOCDF-GC) and OCD Collaborative Genetics Association Studies (OCGAS), 2018. Revealing the complex genetic architecture of obsessive–compulsive disorder using meta-analysis. Mol. Psychiatry 23, 1181–1188. https://doi.org/10.1038/mp.2017.154

Jaffe, A.E., Straub, R.E., Shin, J.H., Tao, R., Gao, Y., Collado-Torres, L., Kam-Thong, T., Xi, H.S., Quan, J., Chen, Q., Colantuoni, C., Ulrich, W.S., Maher, B.J., Deep-Soboslay, A., Cross, A.J., Brandon, N.J., Leek, J.T., Hyde, T.M., Kleinman, J.E., Weinberger, D.R., 2018. Developmental and genetic regulation of the human cortex transcriptome illuminate schizophrenia pathogenesis. Nat. Neurosci. 21, 1117–1125. https://doi.org/10.1038/s41593-018-0197-y

Judd, N., Sauce, B., Wiedenhoeft, J., Tromp, J., Chaarani, B., Schliep, A., van Noort, B., Penttilä, J., Grimmer, Y., Insensee, C., Becker, A., Banaschewski, T., Bokde, A.L.W., Quinlan, E.B., Desrivières, S., Flor, H., Grigis, A., Gowland, P., Heinz, A., Ittermann, B., Martinot, J.-L., Paillère Martinot, M.-L., Artiges, E., Nees, F., Papadopoulos Orfanos, D., Paus, T., Poustka, L., Hohmann, S., Millenet, S., Fröhner, J.H., Smolka, M.N., Walter, H., Whelan, R., Schumann, G., Garavan, H., Klingberg, T., 2020. Cognitive and brain development is independently influenced by socioeconomic status and polygenic scores for educational attainment. Proc. Natl. Acad. Sci. 117, 12411–12418. https://doi.org/10.1073/pnas.2001228117

Keysers, C., Gazzola, V., Wagenmakers, E.-J., 2020. Using Bayes factor hypothesis testing in neuroscience to establish evidence of absence. Nat. Neurosci. 23, 788–799. https://doi.org/10.1038/s41593-020-0660-4

Khundrakpam, B., Vainik, U., Gong, J., Al-Sharif, N., Bhutani, N., Kiar, G., Zeighami, Y., Kirschner, M., Luo, C., Dagher, A., Evans, A., 2020. Neural correlates of polygenic risk score for autism spectrum disorders in general population. Brain Commun. fcaa092. https://doi.org/10.1093/braincomms/fcaa092

Kirschner, M., Paquola, C., Khundrakpam, B.S., Vainik, U., Bhutani, N., Benazir-Hodzic-Santor Al-Sharif, N.B., Misic, B., Bernhardt, B., Evans, A.C., Dagher, A., 2021. Schizophrenia polygenic risk during typical development reflects multiscale cortical organization (preprint). Neuroscience. https://doi.org/10.1101/2021.06.13.448243

Lam, M., Awasthi, S., Watson, H.J., Goldstein, J., Panagiotaropoulou, G., Trubetskoy, V., Karlsson, R., Frei, O., Fan, C.-C., De Witte, W., Mota, N.R., Mullins, N., Brügger, K., Lee, S.H., Wray, N.R., Skarabis, N., Huang, H., Neale, B., Daly, M.J., Mattheisen, M., Walters, R., Ripke, S., 2019. RICOPILI: Rapid Imputation for COnsortias PIpeLIne. Bioinformatics btz633. https://doi.org/10.1093/bioinformatics/btz633

Larsen, B., Luna, B., 2018. Adolescence as a neurobiological critical period for the development of higher-order cognition. Neurosci. Biobehav. Rev. 94, 179–195. https://doi.org/10.1016/j.neubiorev.2018.09.005

Lazari, A., Salvan, P., Cottaar, M., Papp, D., Jens van der Werf, O., Johnstone, A., Sanders, Z.-B., Sampaio-Baptista, C., Eichert, N., Miyamoto, K., Winkler, A., Callaghan, M.F., Nichols, T.E., Stagg, C.J., Rushworth, M.F.S., Verhagen, L., Johansen-Berg, H., 2021. Reassessing associations between white matter and behaviour with multimodal microstructural imaging. Cortex 145, 187–200. https://doi.org/10.1016/j.cortex.2021.08.017

Lee, J.J., Wedow, R., Okbay, A., Kong, E., Maghzian, O., Zacher, M., Nguyen-Viet, T.A., Bowers, P., Sidorenko, J., Linnér, R.K., Fontana, M.A., Kundu, T., Lee, C., Li, H., Li, R., Royer, R., Timshel, P.N., Walters, R.K., Willoughby, E.A., Yengo, L., Alver, M., Bao, Y., Clark, D.W., Day, F.R., Furlotte, N.A., Joshi, P.K., Kemper, K.E., Kleinman, A., Langenberg, C., Mägi, R., Trampush, J.W., Verma, S.S., Wu, Y., Lam, M., Zhao, J.H., Zheng, Z., Boardman, J.D., Campbell, H., Freese, J., Harris, K.M., Hayward, C., Herd, P., Kumari, M., Lencz, T., Luan, J., Malhotra, A.K., Metspalu, A., Milani, L., Ong, K.K., Perry, J.R.B., Porteous, D.J., Ritchie, M.D., Smart, M.C., Smith, B.H., Tung, J.Y., Wareham, N.J., Wilson, J.F., Beauchamp, J.P., Conley, D.C., Esko, T., Lehrer, S.F., Magnusson, P.K.E., Oskarsson, S., Pers, T.H., Robinson, M.R., Thom, K., Watson, C., Chabris, C.F., Meyer, M.N., Laibson, D.I., Yang, J., Johannesson, M., Koellinger, P.D., Turley, P., Visscher, P.M., Benjamin, D.J., Cesarini, D., 2018. Gene discovery and polygenic prediction from a genome-wide association study of educational attainment in 1.1 million individuals. Nat. Genet. 50, 1112–1121. https://doi.org/10.1038/s41588-018-0147-3

Lee, P.H., Anttila, V., Won, H., Feng, Y.-C.A., Rosenthal, J., Zhu, Z., Tucker-Drob, E.M., Nivard, M.G., Grotzinger, A.D., Posthuma, D., Wang, M.M.-J., Yu, D., Stahl, E.A., Walters, R.K., Anney, R.J.L., Duncan, L.E., Ge, T., Adolfsson, R., Banaschewski, T., Belangero, S., Cook, E.H., Coppola, G., Derks, E.M., Hoekstra, P.J., Kaprio, J., Keski-Rahkonen, A., Kirov, G., Kranzler, H.R., Luykx, J.J., Rohde, L.A., Zai, C.C., Agerbo, E., Arranz, M.J., Asherson, P., Bækvad-Hansen, M., Baldursson, G., Bellgrove, M., Belliveau, R.A., Buitelaar, J., Burton, C.L., Bybjerg-Grauholm, J., Casas, M., Cerrato, F., Chambert, K., Churchhouse, C., Cormand, B., Crosbie, J., Dalsgaard, S., Demontis, D., Doyle, A.E., Dumont, A., Elia, J., Grove, J., Gudmundsson, O.O., Haavik, J., Hakonarson, H., Hansen, C.S., Hartman, C.A., Hawi, Z., Hervás, A., Hougaard, D.M., Howrigan, D.P., Huang, H., Kuntsi, J., Langley, K., Lesch, K.-P., Leung, P.W.L., Loo, S.K., Martin, J., Martin, A.R., McGough, J.J., Medland, S.E., Moran, J.L., Mors, O., Mortensen, P.B., Oades, R.D., Palmer, D.S., Pedersen, C.B., Pedersen, M.G., Peters, T., Poterba, T., Poulsen, J.B., Ramos-Quiroga, J.A., Reif, A., Ribasés, M., Rothenberger, A., Rovira, P., Sánchez-Mora, C., Satterstrom, F.K., Schachar, R., Artigas, M.S., Steinberg, S., Stefansson, H., Turley, P., Walters, G.B., Werge, T., Zayats, T., Arking, D.E., Bettella, F., Buxbaum, J.D., Christensen, J.H., Collins, R.L., Coon, H., De Rubeis, S., Delorme, R., Grice, D.E., Hansen, T.F., Holmans, P.A., Hope, S., Hultman, C.M., Klei, L., Ladd-Acosta, C., Magnusson, P., Nærland, T., Nyegaard, M., Pinto, D., Qvist, P., Rehnström, K., Reichenberg, A., Reichert, J., Roeder, K., Rouleau, G.A., Saemundsen, E., Sanders, S.J., Sandin, S., St Pourcain, B., Stefansson, K., Sutcliffe, J.S., Talkowski, M.E., Weiss, L.A., Willsey, A.J., Agartz, I., Akil, H., Albani, D., Alda, M., Als, T.D., Anjorin, A., Backlund, L., Bass, N., Bauer, M., Baune, B.T., Bellivier, F., Bergen, S.E., Berrettini, W.H., Biernacka, J.M., Blackwood, D.H.R., Bøen, E., Budde, M., Bunney, W., Burmeister, M., Byerley, W., Byrne, E.M., Cichon, S., Clarke, T.-K., Coleman, J.R.I., Craddock, N., Curtis, D., Czerski, P.M., Dale, A.M., Dalkner, N., Dannlowski, U., Degenhardt, F., Di Florio, A., Elvsåshagen, T., Etain, B., Fischer, S.B., Forstner, A.J., Forty, L., Frank, J., Frye, M., Fullerton, J.M., Gade, K., Gaspar, H.A., Gershon, E.S., Gill, M., Goes, F.S., Gordon, S.D., Gordon-Smith, K., Green, M.J., Greenwood, T.A., Grigoroiu-Serbanescu, M., Guzman-Parra, J., Hauser, J., Hautzinger, M., Heilbronner, U., Herms, S., Hoffmann, P., Holland, D., Jamain, S., Jones, I., Jones, L.A., Kandaswamy, R., Kelsoe, J.R., Kennedy, J.L., Joachim, O.K., Kittel-Schneider, S., Kogevinas, M., Koller, A.C., Lavebratt, C., Lewis, C.M., Li, Q.S., Lissowska, J., Loohuis, L.M.O., Lucae, S., Maaser, A., Malt, U.F., Martin, N.G., Martinsson, L., McElroy, S.L., McMahon, F.J., McQuillin, A., Melle, I., Metspalu, A., Millischer, V., Mitchell, P.B., Montgomery, G.W., Morken, G., Morris, D.W., Müller-Myhsok, B., Mullins, N., Myers, R.M., Nievergelt, C.M., Nordentoft, M., Adolfsson, A.N., Nöthen, M.M., Ophoff, R.A., Owen, M.J., Paciga, S.A., Pato, C.N., Pato, M.T., Perlis, R.H., Perry, A., Potash, J.B., Reinbold, C.S., Rietschel, M., Rivera, M., Roberson, M., Schalling, M., Schofield, P.R., Schulze, T.G., Scott, L.J., Serretti, A., Sigurdsson, E., Smeland, O.B., Stordal, E., Streit, F., Strohmaier, J., Thorgeirsson, T.E., Treutlein, J., Turecki, G., Vaaler, A.E., Vieta, E., Vincent, J.B., Wang, Y., Witt, S.H., Zandi, P., Adan, R.A.H., Alfredsson, L., Ando, T., Aschauer, H., Baker, J.H., Bencko, V., Bergen, A.W., Birgegård, A., Perica, V.B., Brandt, H., Burghardt, R., Carlberg, L., Cassina, M., Clementi, M., Courtet, P., Crawford, S., Crow, S., Crowley, J.J., Danner, U.N., Davis, O.S.P., Degortes, D., DeSocio, J.E., Dick, D.M., Dina, C., Docampo, E., Egberts, K., Ehrlich, S., Espeseth, T., Fernández-Aranda, F., Fichter, M.M., Foretova, L., Forzan, M., Gambaro, G., Giegling, I., Gonidakis, F., Gorwood, P., Mayora, M.G., Guo, Y., Halmi, K.A., Hatzikotoulas, K., Hebebrand, J., Helder, S.G., Herpertz-Dahlmann, B., Herzog, W., Hinney, A., Imgart, H., Jiménez-Murcia, S., Johnson, C., Jordan, J., Julià, A., Kaminská, D., Karhunen, L., Karwautz, A., Kas, M.J.H., Kaye, W.H., Kennedy, M.A., Kim, Y.-R., Klareskog, L., Klump, K.L., Knudsen, G.P.S., Landén, M., Le Hellard, S., Levitan, R.D., Li, D., Lichtenstein, P., Maj, M., Marsal, S., McDevitt, S., Mitchell, J., Monteleone, P., Monteleone, A.M., Munn-Chernoff, M.A., Nacmias, B., Navratilova, M., O’Toole, J.K., Padyukov, L., Pantel, J., Papezova, H., Rabionet, R., Raevuori, A., Ramoz, N., Reichborn-Kjennerud, T., Ricca, V., Roberts, M., Rujescu, D., Rybakowski, F., Scherag, A., Schmidt, U., Seitz, J., Slachtova, L., Slof-Op’t Landt, M.C.T., Slopien, A., Sorbi, S., Southam, L., Strober, M., Tortorella, A., Tozzi, F., Treasure, J., Tziouvas, K., van Elburg, A.A., Wade, T.D., Wagner, G., Walton, E., Watson, H.J., Wichmann, H.-E., Woodside, D.B., Zeggini, E., Zerwas, S., Zipfel, S., Adams, M.J., Andlauer, T.F.M., Berger, K., Binder, E.B., Boomsma, D.I., Castelao, E., Colodro-Conde, L., Direk, N., Docherty, A.R., Domenici, E., Domschke, K., Dunn, E.C., Foo, J.C., de. Geus, E.J.C., Grabe, H.J., Hamilton, S.P., Horn, C., Hottenga, J.-J., Howard, D., Ising, M., Kloiber, S., Levinson, D.F., Lewis, G., Magnusson, P.K.E., Mbarek, H., Middeldorp, C.M., Mostafavi, S., Nyholt, D.R., Penninx, B.WJH., Peterson, R.E., Pistis, G., Porteous, D.J., Preisig, M., Quiroz, J.A., Schaefer, C., Schulte, E.C., Shi, J., Smith, D.J., Thomson, P.A., Tiemeier, H., Uher, R., van der Auwera, S., Weissman, M.M., Alexander, M., Begemann, M., Bramon, E., Buccola, N.G., Cairns, M.J., Campion, D., Carr, V.J., Cloninger, C.R., Cohen, D., Collier, D.A., Corvin, A., DeLisi, L.E., Donohoe, G., Dudbridge, F., Duan, J., Freedman, R., Gejman, P.V., Golimbet, V., Godard, S., Ehrenreich, H., Hartmann, A.M., Henskens, F.A., Ikeda, M., Iwata, N., Jablensky, A.V., Joa, I., Jönsson, E.G., Kelly, B.J., Knight, J., Konte, B., Laurent-Levinson, C., Lee, J., Lencz, T., Lerer, B., Loughland, C.M., Malhotra, A.K., Mallet, J., McDonald, C., Mitjans, M., Mowry, B.J., Murphy, K.C., Murray, R.M., O’Neill, F.A., Oh, S.-Y., Palotie, A., Pantelis, C., Pulver, A.E., Petryshen, T.L., Quested, D.J., Riley, B., Sanders, A.R., Schall, U., Schwab, S.G., Scott, R.J., Sham, P.C., Silverman, J.M., Sim, K., Steixner, A.A., Tooney, P.A., van Os, J., Vawter, M.P., Walsh, D., Weiser, M., Wildenauer, D.B., Williams, N.M., Wormley, B.K., Zhang, F., Androutsos, C., Arnold, P.D., Barr, C.L., Barta, C., Bey, K., Bienvenu, O.J., Black, D.W., Brown, L.W., Budman, C., Cath, D., Cheon, K.-A., Ciullo, V., Coffey, B.J., Cusi, D., Davis, L.K., Denys, D., Depienne, C., Dietrich, A., Eapen, V., Falkai, P., Fernandez, T.V., Garcia-Delgar, B., Geller, D.A., Gilbert, D.L., Grados, M.A., Greenberg, E., Grünblatt, E., Hagstrøm, J., Hanna, G.L., Hartmann, A., Hedderly, T., Heiman, G.A., Heyman, I., Hong, H.J., Huang, A., Huyser, C., Ibanez-Gomez, L., Khramtsova, E.A., Kim, Y.K., Kim, Y.-S., King, R.A., Koh, Y.-J., Konstantinidis, A., Kook, S., Kuperman, S., Leventhal, B.L., Lochner, C., Ludolph, A.G., Madruga-Garrido, M., Malaty, I., Maras, A., McCracken, J.T., Meijer, I.A., Mir, P., Morer, A., Müller-Vahl, K.R., Münchau, A., Murphy, T.L., Naarden, A., Nagy, P., Nestadt, G., Nestadt, P.S., Nicolini, H., Nurmi, E.L., Okun, M.S., Paschou, P., Piras, Fabrizio, Piras, Federica, Pittenger, C., Plessen, K.J., Richter, M.A., Rizzo, R., Robertson, M., Roessner, V., Ruhrmann, S., Samuels, J.F., Sandor, P., Schlögelhofer, M., Shin, E.-Y., Singer, H., Song, D.-H., Song, J., Spalletta, G., Stein, D.J., Stewart, S.E., Storch, E.A., Stranger, B., Stuhrmann, M., Tarnok, Z., Tischfield, J.A., Tübing, J., Visscher, F., Vulink, N., Wagner, M., Walitza, S., Wanderer, S., Woods, M., Worbe, Y., Zai, G., Zinner, S.H., Sullivan, P.F., Franke, B., Daly, M.J., Bulik, C.M., Lewis, C.M., McIntosh, A.M., O’Donovan, M.C., Zheutlin, A., Andreassen, O.A., Børglum, A.D., Breen, G., Edenberg, H.J., Fanous, A.H., Faraone, S.V., Gelernter, J., Mathews, C.A., Mattheisen, M., Mitchell, K.S., Neale, M.C., Nurnberger, J.I., Ripke, S., Santangelo, S.L., Scharf, J.M., Stein, M.B., Thornton, L.M., Walters, J.T.R., Wray, N.R., Geschwind, D.H., Neale, B.M., Kendler, K.S., Smoller, J.W., 2019. Genomic Relationships, Novel Loci, and Pleiotropic Mechanisms across Eight Psychiatric Disorders. Cell 179, 1469-1482.e11. https://doi.org/10.1016/j.cell.2019.11.020

Lenroot, R.K., Schmitt, J.E., Ordaz, S.J., Wallace, G.L., Neale, M.C., Lerch, J.P., Kendler, K.S., Evans, A.C., Giedd, J.N., 2009. Differences in genetic and environmental influences on the human cerebral cortex associated with development during childhood and adolescence. Hum. Brain Mapp. 30, 163–174. https://doi.org/10.1002/hbm.20494

Li, M., Santpere, G., Imamura Kawasawa, Y., Evgrafov, O.V., Gulden, F.O., Pochareddy, S., Sunkin, S.M., Li, Zhen, Shin, Y., Zhu, Y., Sousa, A.M.M., Werling, D.M., Kitchen, R.R., Kang, H.J., Pletikos, M., Choi, J., Muchnik, S., Xu, X., Wang, D., Lorente-Galdos, B., Liu, S., Giusti-Rodríguez, P., Won, H., de Leeuw, C.A., Pardiñas, A.F., BrainSpan Consortium, PsychENCODE Consortium, PsychENCODE Developmental Subgroup, Hu, M., Jin, F., Li, Y., Owen, M.J., O’Donovan, M.C., Walters, J.T.R., Posthuma, D., Reimers, M.A., Levitt, P., Weinberger, D.R. Hyde, Thomas M., Kleinman, J.E., Geschwind, D.H., Hawrylycz, M.J., State, M.W., Sanders, S.J., Sullivan, P.F., Gerstein, M.B., Lein, E.S., Knowles, J.A., Sestan, N., Willsey, A.J., Oldre, A., Szafer, A., Camarena, A., Cherskov, A., Charney, A.W., Abyzov, A., Kozlenkov, A., Safi, A., Jones, A.R., Ashley-Koch, A.E., Ebbert, A., Price, A.J., Sekijima, A., Kefi, A., Bernard, A., Amiri, A., Sboner, A., Clark, A., Jaffe, A.E., Tebbenkamp, A.T.N., Sodt, A.J., Guillozet-Bongaarts, A.L., Nairn, A.C., Carey, A., Huttner, A., Chervenak, A., Szekely, A., Shieh, A.W., Harmanci, A., Lipska, B.K., Carlyle, B.C., Gregor, B.W., Kassim, B.S., Sheppard, B., Bichsel, C., Hahn, C.-G., Lee, C.-K., Chen, C., Kuan, C.L., Dang, C., Lau, C., Cuhaciyan, C., Armoskus, C., Mason, C.E., Liu, C., Slaughterbeck, C.R., Bennet, C., Pinto, D., Polioudakis, D., Franjic, D., Miller, D.J., Bertagnolli, D., Lewis, D.A., Feng, D., Sandman, D., Clarke, D., Williams, D., DelValle, D., Fitzgerald, D., Shen, E.H., Flatow, E., Zharovsky, E., Burke, E.E., Olson, E., Fulfs, E., Mattei, E., Hadjimichael, E., Deelman, E., Navarro, F.C.P., Wu, F., Lee, F., Cheng, F., Goes, F.S., Vaccarino, F.M., Liu, F., Hoffman, G.E., Gürsoy, G., Gee, G., Mehta, G., Coppola, G., Giase, G., Sedmak, G., Johnson, G.D., Wray, G.A., Crawford, G.E., Gu, G., van Bakel, H., Witt, H., Yoon, H.J., Pratt, H., Zhao, H., Glass, I.A., Huey, J., Arnold, J., Noonan, J.P., Bendl, J., Jochim, J.M., Goldy, J., Herstein, J., Wiseman, J.R., Miller, J.A., Mariani, J., Stoll, J., Moore, J., Szatkiewicz, J., Leng, J., Zhang, J., Parente, J., Rozowsky, J., Fullard, J.F., Hohmann, J.G., Morris, J., Phillips, J.W., Warrell, J., Shin, J.H., An, J.-Y., Belmont, J., Nyhus, J., Pendergraft, J., Bryois, J., Roll, K., Grennan, K.S., Aiona, K., White, K.P., Aldinger, K.A., Smith, K.A., Girdhar, K., Brouner, K., Mangravite, L.M., Brown, L., Collado-Torres, L., Cheng, L., Gourley, L., Song, L., Ubieta, L.D.L.T., Habegger, L., Ng, L., Hauberg, M.E., Onorati, M., Webster, M.J., Kundakovic, M., Skarica, M., Reimers, M., Johnson, M.B., Chen, M.M., Garrett, M.E., Sarreal, M., Reding, M., Gu, M., Peters, M.A., Fisher, M., Gandal, M.J., Purcaro, M., Smith, M., Brown, Miguel, Shibata, M., Brown, Mimi, Xu, M., Yang, M., Ray, M., Shapovalova, N.V., Francoeur, N., Sjoquist, N., Mastan, N., Kaur, N., Parikshak, N., Mosqueda, N.F., Ngo, N.-K., Dee, N., Ivanov, N.A., Devillers, O., Roussos, P., Parker, P.D., Manser, P., Wohnoutka, P., Farnham, P.J., Zandi, P., Emani, P.S., Dalley, R.A., Mayani, R., Tao, R., Gittin, R., Straub, R.E., Lifton, R.P., Jacobov, R., Howard, R.E., Park, R.B., Dai, R., Abramowicz, S., Akbarian, S., Schreiner, S., Ma, S., Parry, S.E., Shapouri, S., Weissman, S., Caldejon, S., Mane, S., Ding, S.-L., Scuderi, S., Dracheva, S., Butler, S., Lisgo, S.N., Rhie, S.K., Lindsay, S., Datta, S., Souaiaia, T., Roychowdhury, T., Gomez, T., Naluai-Cecchini, T., Beach, T.G., Goodman, T., Gao, T., Dolbeare, T.A., Fliss, T., Reddy, T.E., Chen, T. Hyde, Tom M., Brunetti, T., Lemon, T.A., Desta, T., Borrman, T., Haroutunian, V., Spitsyna, V.N., Swarup, V., Shi, X., Jiang, Yan, Xia, Y., Chen, Y.-H., Jiang, Yi, Wang, Y., Chae, Y., Yang, Y.T., Kim, Y., Riley, Z.L., Krsnik, Z., Deng, Z., Weng, Z., Lin, Z., Li, Zhuo, 2018. Integrative functional genomic analysis of human brain development and neuropsychiatric risks. Science 362, eaat7615. https://doi.org/10.1126/science.aat7615

Marco, E.M., Macrì, S., Laviola, G., 2011. Critical age windows for neurodevelopmental psychiatric disorders: evidence from animal models. Neurotox. Res. 19, 286–307. https://doi.org/10.1007/s12640-010-9205-z

McIntosh, A.R., 2021. Comparison of Canonical Correlation and Partial Least Squares analyses of simulated and empirical data. ArXiv210706867 Stat.

Miller, K.L., Alfaro-Almagro, F., Bangerter, N.K., Thomas, D.L., Yacoub, E., Xu, J., Bartsch, A.J., Jbabdi, S., Sotiropoulos, S.N., Andersson, J.L.R., Griffanti, L., Douaud, G., Okell, T.W., Weale, P., Dragonu, I., Garratt, S., Hudson, S., Collins, R., Jenkinson, M., Matthews, P.M., Smith, S.M., 2016. Multimodal population brain imaging in the UK Biobank prospective epidemiological study. Nat. Neurosci. 19, 1523–1536. https://doi.org/10.1038/nn.4393

Mowinckel, A.M., Vidal-Piñeiro, D., 2020. Visualization of Brain Statistics With R Packages ggseg and ggseg3d. Adv. Methods Pract. Psychol. Sci. 3, 466–483. https://doi.org/10.1177/2515245920928009

Natu, V.S., Gomez, J., Barnett, M., Jeska, B., Kirilina, E., Jaeger, C., Zhen, Z., Cox, S., Weiner, K.S., Weiskopf, N., Grill-Spector, K., 2019. Apparent thinning of human visual cortex during childhood is associated with myelination. Proc. Natl. Acad. Sci. 116, 20750– 20759. https://doi.org/10.1073/pnas.1904931116

Ni, G., Zeng, J., Revez, J.A., Wang, Ying, Zheng, Z., Ge, T., Restuadi, R., Kiewa, J., Nyholt, D.R., Coleman, J.R.I., Smoller, J.W., Yang, J., Visscher, P.M., Wray, N.R., Ripke, S., Neale, B.M., Corvin, A., Walters, J.T.R., Farh, K.-H., Holmans, P.A., Lee, P., Bulik-Sullivan, B., Collier, D.A., Huang, H., Pers, T.H., Agartz, I., Agerbo, E., Albus, M., Alexander, M., Amin, F., Bacanu, S.A., Begemann, M., Belliveau, R.A., Bene, J., Bergen, S.E., Bevilacqua, E., Bigdeli, T.B., Black, D.W., Bruggeman, R., Buccola, N.G., Buckner, R.L., Byerley, W., Cahn, W., Cai, G., Campion, D., Cantor, R.M., Carr, V.J., Carrera, N., Catts, S.V., Chambert, K.D., Chan, R.C.K., Chen, R.Y.L., Chen, E.Y.H., Cheng, W., Cheung, E.F.C., Chong, S.A., Cloninger, C.R., Cohen, D., Cohen, N., Cormican, P., Craddock, N., Crowley, J.J., Davidson, M., Davis, K.L., Degenhardt, F., Del Favero, J., Demontis, D., Dikeos, D., Dinan, T., Djurovic, S., Donohoe, G., Drapeau, E., Duan, J., Dudbridge, F., Durmishi, N., Eichhammer, P., Eriksson, J., Escott-Price, V., Essioux, L., Fanous, A.H., Farrell, M.S., Frank, J., Franke, L., Freedman, R., Freimer, N.B., Friedl, M., Friedman, J.I., Fromer, M., Genovese, G., Georgieva, L., Giegling, I., Giusti-Rodríguez, P., Godard, S., Goldstein, J.I., Golimbet, V., Gopal, S., Gratten, J., de Haan, L., Hammer, C., Hamshere, M.L., Hansen, M., Hansen, T., Haroutunian, V., Hartmann, A.M., Henskens, F.A., Herms, S., Hirschhorn, J.N., Hoffmann, P., Hofman, A., Hollegaard, M.V., Hougaard, D.M., Ikeda, M., Joa, I., Julià, A., Kahn, R.S., Kalaydjieva, L., Karachanak-Yankova, S., Karjalainen, J., Kavanagh, D., Keller, M.C., Kennedy, J.L., Khrunin, A., Kim, Y., Klovins, J., Knowles, J.A., Konte, B., Kucinskas, V., Kucinskiene, Z.A., Kuzelova-Ptackova, H., Kähler, A.K., Laurent, C., Lee, J., Lee, S.H., Legge, S.E., Lerer, B., Li, M., Li, T., Liang, K.-Y., Lieberman, J., Limborska, S., Loughland, C.M., Lubinski, J., Lönnqvist, J., Macek, M., Magnusson, P.K.E., Maher, B.S., Maier, W., Mallet, J., Marsal, S., Mattheisen, M., Mattingsdal, M., McCarley, R.W., McDonald, C., McIntosh, A.M., Meier, S., Meijer, C.J., Melegh, B., Melle, I., Mesholam-Gately, R.I., Metspalu, A., Michie, P.T., Milani, L., Milanova, V., Mokrab, Y., Morris, D.W., Mors, O., Murphy, K.C., Murray, R.M., Myin-Germeys, I., Müller-Myhsok, B., Nelis, M., Nenadic, I., Nertney, D.A., Nestadt, G., Nicodemus, K.K., Nikitina-Zake, L., Nisenbaum, L., Nordin, A., O’Callaghan, E., O’Dushlaine, C., O’Neill, F.A., Oh, S.-Y., Olincy, A., Olsen, L., Van Os, J., International Consortium, P.E., Pantelis, C., Papadimitriou, G.N., Papiol, S., Parkhomenko, E., Pato, M.T., Paunio, T., Pejovic-Milovancevic, M., Perkins, D.O., Pietiläinen, O., Pimm, J., Pocklington, A.J., Powell, J., Price, A., Pulver, A.E., Purcell, S.M., Quested, D., Rasmussen, H.B., Reichenberg, A., Reimers, M.A., Richards, A.L., Roffman, J.L., Roussos, P., Ruderfer, D.M., Salomaa, V., Sanders, A.R., Schall, U., Schubert, C.R., Schulze, T.G., Schwab, S.G., Scolnick, E.M., Scott, R.J., Seidman, L.J., Shi, J., Sigurdsson, E., Silagadze, T., Silverman, J.M., Sim, K., Slominsky, P., Smoller, J.W., So, H.-C., Spencer, C.C.A., Stahl, E.A., Stefansson, H., Steinberg, S., Stogmann, E., Straub, R.E., Strengman, E., Strohmaier, J., Stroup, T.S., Subramaniam, M., Suvisaari, J., Svrakic, D.M., Szatkiewicz, J.P., Söderman, E., Thirumalai, S., Toncheva, D., Tosato, S., Veijola, J., Waddington, J., Walsh, D., Wang, D., Wang, Q., Webb, B.T., Weiser, M., Wildenauer, D.B., Williams, N.M., Williams, S., Witt, S.H., Wolen, A.R., Wong, E.H.M., Wormley, B.K., Xi, Hualin Simon, Zai, C.C., Zheng, X., Zimprich, F., Wray, N.R., Stefansson, K., Visscher, P.M., Case-Control Consortium, W.T., Adolfsson, R., Andreassen, O.A., Blackwood, D.H.R., Bramon, E., Buxbaum, J.D., Børglum, A.D., Cichon, S., Darvasi, A., Domenici, E., Ehrenreich, H., Esko, T., Gejman, P.V., Gill, M., Gurling, H., Hultman, C.M., Iwata, N., Jablensky, A.V., Jönsson, E.G., Kendler, K.S., Kirov, G., Knight, J., Lencz, T., Levinson, D.F., Li, Q.S., Liu, J., Malhotra, A.K., McCarroll, S.A., McQuillin, A., Moran, J.L., Mortensen Preben B., Mowry, B.J., Nöthen, M.M., Ophoff, R.A., Owen, M.J., Palotie, A., Pato, C.N., Petryshen, T.L., Posthuma, D., Rietschel, M., Riley, B.P., Rujescu, D., Sham, P.C., Sklar, P., St Clair, D., Weinberger, D.R., Wendland, J.R., Werge, T., Daly, M.J., Sullivan, P.F., O’Donovan, M.C., Wray, N.R., Ripke, S., Mattheisen, M., Trzaskowski, M., Byrne, E.M., Abdellaoui, A., Adams, M.J., Agerbo, E., Air, T.M., Andlauer, T.F.M., Bacanu, S.-A., Bækvad-Hansen, M., Beekman, A.T.F., Bigdeli, T.B., Binder, E.B., Bryois, J., Buttenschøn, H.N., Bybjerg-Grauholm, J., Cai, N., Castelao, E., Christensen, J.H., Clarke, T.-K., Coleman, J.R.I., Colodro-Conde, L., Couvy-Duchesne, B., Craddock, N., Crawford, G.E., Davies, G., Deary, I.J., Degenhardt, F., Derks, E.M., Direk, N., Dolan, C.V., Dunn, E.C., Eley, T.C., Escott-Price, V., Hassan Kiadeh, F.F., Finucane, H.K., Foo, J.C., Forstner, A.J., Frank, J., Gaspar, H.A., Gill, M., Goes, F.S., Gordon, S.D., Grove, J., Hall, L.S., Hansen, C.S., Hansen, T.F., Herms, S., Hickie, I.B., Hoffmann, P., Homuth, G., Horn, C., Hottenga, J.-J., Hougaard, D.M., Howard, D.M., Ising, M., Jansen, R., Jones, I., Jones, L.A., Jorgenson, E., Knowles, J.A., Kohane, I.S., Kraft, J., Kretzschmar, W.W., Kutalik, Z., Li, Y., Lind, P.A., MacIntyre, D.J., MacKinnon, D.F., Maier, R.M., Maier, W., Marchini, J., Mbarek, H., McGrath, P., McGuffin, P., Medland, S.E., Mehta, D., Middeldorp, C.M., Mihailov, E., Milaneschi, Y., Milani, L., Mondimore, F.M., Montgomery, G.W., Mostafavi, S., Mullins, N., Nauck, M., Ng, B., Nivard, M.G., Nyholt, D.R., O’Reilly, P.F., Oskarsson, H., Owen, M.J., Painter, J.N., Pedersen, C.B., Pedersen, M.G., Peterson, R.E., Peyrot, W.J., Pistis, G., Posthuma, D., Quiroz, J.A., Qvist, P., Rice, J.P., Riley, B.P., Rivera, M., Mirza, S.S., Schoevers, R., Schulte, E.C., Shen, L., Shi, J., Shyn, S.I., Sigurdsson, E., Sinnamon, G.C.B., Smit, J.H., Smith, D.J., Stefansson, H., Steinberg, S., Streit, F., Strohmaier, J., Tansey, K.E., Teismann, H., Teumer, A., Thompson, W., Thomson, P.A., Thorgeirsson, T.E., Traylor, M., Treutlein, J., Trubetskoy, V., Uitterlinden, A.G., Umbricht, D., Van der Auwera, S., van Hemert, A.M., Viktorin, A., Visscher, P.M., Wang, Yunpeng, Webb, B.T., Weinsheimer, S.M., Wellmann, J., Willemsen, G., Witt, S.H., Wu, Y. Xi, Hualin S., Yang, J., Zhang, F., Arolt, V., Baune, B.T., Berger, K., Boomsma, D.I., Cichon, S., Dannlowski, U., de Geus, E.J.C., DePaulo, J.R., Domenici, E., Domschke, K., Esko, T., Grabe, H.J., Hamilton, S.P., Hayward, C., Heath, A.C., Kendler, K.S., Kloiber, S., Lewis, G., Li, Q.S., Lucae, S., Madden, P.A.F., Magnusson, P.K., Martin, N.G., McIntosh, A.M., Metspalu, A., Mors, O., Mortensen, Preben Bo, Müller-Myhsok, B., Nordentoft, M., Nöthen, M.M., O’Donovan, M.C., Paciga, S.A., Pedersen, N.L., 2021. A Comparison of Ten Polygenic Score Methods for Psychiatric Disorders Applied Across Multiple Cohorts. Biol. Psychiatry 90, 611–620. https://doi.org/10.1016/j.biopsych.2021.04.018

Norbom, L.B., Ferschmann, L., Parker, N., Agartz, I., Andreassen, O.A., Paus, T., Westlye, L.T., Tamnes, C.K., 2021. New insights into the dynamic development of the cerebral cortex in childhood and adolescence: Integrating macro- and microstructural MRI findings. Prog. Neurobiol. 204, 102109. https://doi.org/10.1016/j.pneurobio.2021.102109

Palmer, C.E., Pecheva, D., Iversen, J.R., Hagler, D.J., Sugrue, L., Nedelec, P., Fan, C.C., Thompson, W.K., Jernigan, T.L., Dale, A.M., 2021. Microstructural development from 9-14 years: evidence from the ABCD Study. Dev. Cogn. Neurosci. 101044. https://doi.org/10.1016/j.dcn.2021.101044

Paulus, M.P., Thompson, W.K., 2019. The Challenges and Opportunities of Small Effects: The New Normal in Academic Psychiatry. JAMA Psychiatry 76, 353. https://doi.org/10.1001/jamapsychiatry.2018.4540

Purcell, S., Neale, B., Todd-Brown, K., Thomas, L., Ferreira, M.A.R., Bender, D., Maller, J., Sklar, P., de Bakker, P.I.W., Daly, M.J., Sham, P.C., 2007. PLINK: A Tool Set for Whole-Genome Association and Population-Based Linkage Analyses. Am. J. Hum. Genet. 81, 559–575. https://doi.org/10.1086/519795

Quintana, D.S., Williams, D.R., 2018. Bayesian alternatives for common null-hypothesis significance tests in psychiatry: a non-technical guide using JASP. BMC Psychiatry 18, 178. https://doi.org/10.1186/s12888-018-1761-4

Ripke, S., Walters, J.T., O’Donovan, M.C., 2020. Mapping genomic loci prioritises genes and implicates synaptic biology in schizophrenia (preprint). Genetic and Genomic Medicine. https://doi.org/10.1101/2020.09.12.20192922

Ritchie, S.J., Quinlan, E.B., Banaschewski, T., Bokde, A.L.W., Desrivieres, S., Flor, H., Frouin, V., Garavan, H., Gowland, P., Heinz, A., Ittermann, B., Martinot, J.-L., Martinot, M.-L.P., Nees, F., Papadopoulos Orfanos, D., Paus, T., Poustka, L., Hohmann, S., Millenet, S., Fröhner, J., Smolka, M.N., Walter, H., Whelan, R., Schumann, G., 2019. Neuroimaging and genetic correlates of cognitive ability and cognitive development in adolescence (preprint). PsyArXiv. https://doi.org/10.31234/osf.io/8pwd6

Rokicki, J., Wolfers, T., Nordhøy, W., Tesli, N., Quintana, D.S., Alnæs, D., Richard, G., Lange, A.G., Lund, M.J., Norbom, L., Agartz, I., Melle, I., Nærland, T., Selbæk, G., Persson, K., Nordvik, J.E., Schwarz, E., Andreassen, O.A., Kaufmann, T., Westlye, L.T., 2021. Multimodal imaging improves brain age prediction and reveals distinct abnormalities in patients with psychiatric and neurological disorders. Hum. Brain Mapp. 42, 1714–1726. https://doi.org/10.1002/hbm.25323

Schmitt, J.E., Neale, M.C., Clasen, L.S., Liu, S., Seidlitz, J., Pritikin, J.N., Chu, A., Wallace, G.L., Lee, N.R., Giedd, J.N., Raznahan, A., 2019. A Comprehensive Quantitative Genetic Analysis of Cerebral Surface Area in Youth. J. Neurosci. Off. J. Soc. Neurosci. 39, 3028–3040. https://doi.org/10.1523/JNEUROSCI.2248-18.2019

Schmitt, J.E., Neale, M.C., Fassassi, B., Perez, J., Lenroot, R.K., Wells, E.M., Giedd, J.N., 2014. The dynamic role of genetics on cortical patterning during childhood and adolescence. Proc. Natl. Acad. Sci. 111, 6774–6779. https://doi.org/10.1073/pnas.1311630111

Schmitt, J.E., Raznahan, A., Liu, S., Neale, M.C., 2020. The genetics of cortical myelination in young adults and its relationships to cerebral surface area, cortical thickness, and intelligence: A magnetic resonance imaging study of twins and families. NeuroImage 206, 116319. https://doi.org/10.1016/j.neuroimage.2019.116319

Schweiger, R., Fisher, E., Rahmani, E., Shenhav, L., Rosset, S., Halperin, E., 2018. Using Stochastic Approximation Techniques to Efficiently Construct Confidence Intervals for Heritability. J. Comput. Biol. 25, 794–808. https://doi.org/10.1089/cmb.2018.0047

Schweiger, R., Fisher, E., Rahmani, E., Shenhav, L., Rosset, S., Halperin, E., 2017. Using Stochastic Approximation Techniques to Efficiently Construct Confidence Intervals for Heritability, in: Sahinalp, S.C. (Ed.), Research in Computational Molecular Biology, Lecture Notes in Computer Science. Springer International Publishing, Cham, pp. 241–256. https://doi.org/10.1007/978-3-319-56970-3_15

Sekar, Aswin, Sekar, A., Bialas, A.R., de Rivera, H., Davis, A., Hammond, T.R., Kamitaki, N., Tooley, K., Presumey, J., Baum, M., Van Doren, V., Genovese, G., Rose, S.A., Handsaker, R.E., Daly, M.J., Carroll, M.C., Stevens, B., McCarroll, S.A., 2016. Schizophrenia risk from complex variation of complement component 4. Nature 530, 177–183. https://doi.org/10.1038/nature16549

Smith, S.M., Douaud, G., Chen, W., Hanayik, T., Alfaro-Almagro, F., Sharp, K., Elliott, L.T., 2020. Enhanced Brain Imaging Genetics in UK Biobank (preprint). Neuroscience. https://doi.org/10.1101/2020.07.27.223545

Smith, S.M., Nichols, T.E., 2018. Statistical Challenges in “Big Data” Human Neuroimaging. Neuron 97, 263–268. https://doi.org/10.1016/j.neuron.2017.12.018

Solmi, M., Radua, J., Olivola, M., Croce, E., Soardo, L., Salazar de Pablo, G., Il Shin, J., Kirkbride, J.B., Jones, P., Kim, J.H., Kim, J.Y., Carvalho, A.F., Seeman, M.V., Correll, C.U., Fusar-Poli, P., 2021. Age at onset of mental disorders worldwide: large-scale meta-analysis of 192 epidemiological studies. Mol. Psychiatry. https://doi.org/10.1038/s41380-021-01161-7

Sotiras, A., Toledo, J.B., Gur, R.E., Gur, R.C., Satterthwaite, T.D., Davatzikos, C., 2017. Patterns of coordinated cortical remodeling during adolescence and their associations with functional specialization and evolutionary expansion. Proc. Natl. Acad. Sci. U. S. A. 114, 3527–3532. https://doi.org/10.1073/pnas.1620928114

Sprooten, E., Franke, B., Greven, C.U., 2021. The P-factor and its genomic and neural equivalents: an integrated perspective. Mol. Psychiatry. https://doi.org/10.1038/s41380-021-01031-2

Stahl, E.A., Breen, G., Forstner, A.J., McQuillin, A., Ripke, S., Trubetskoy, V., Mattheisen, M., Wang, Y., Coleman, J.R.I., Gaspar, H.A., de Leeuw, C.A., Steinberg, S., Pavlides, J.M.W., Trzaskowski, M., Byrne, E.M., Pers, T.H., Holmans, P.A., Richards, A.L., Abbott, L., Agerbo, E., Akil, H., Albani, D., Alliey-Rodriguez, N., Als, T.D., Anjorin, A., Antilla, V., Awasthi, S., Badner, J.A., Bækvad-Hansen, M., Barchas, J.D., Bass, N., Bauer, M., Belliveau, R., Bergen, S.E., Pedersen, C.B., Bøen, E., Boks, M.P., Boocock, J., Budde, M., Bunney, W., Burmeister, M., Bybjerg-Grauholm, J., Byerley, W., Casas, M., Cerrato, F., Cervantes, P., Chambert, K., Charney, A.W., Chen, D., Churchhouse, C., Clarke, T.-K., Coryell, W., Craig, D.W., Cruceanu, C., Curtis, D., Czerski, P.M., Dale, A.M., de Jong, S., Degenhardt, F., Del-Favero, J., DePaulo, J.R., Djurovic, S., Dobbyn, A.L., Dumont, A., Elvsåshagen, T., Escott-Price, V., Fan, C.C., Fischer, S.B., Flickinger, M., Foroud, T.M., Forty, L., Frank, J., Fraser, C., Freimer, N.B., Frisén, L., Gade, K., Gage, D., Garnham, J., Giambartolomei, C., Pedersen, M.G., Goldstein, J., Gordon, S.D., Gordon-Smith, K., Green, E.K., Green, M.J., Greenwood, T.A., Grove, J., Guan, W., Guzman-Parra, J., Hamshere, M.L., Hautzinger, M., Heilbronner, U., Herms, S., Hipolito, M., Hoffmann, P., Holland, D., Huckins, L., Jamain, S., Johnson, J.S., Juréus, A., Kandaswamy, R., Karlsson, R., Kennedy, J.L., Kittel-Schneider, S., Knowles, J.A., Kogevinas, M., Koller, A.C., Kupka, R., Lavebratt, C., Lawrence, J., Lawson, W.B., Leber, M., Lee, P.H., Levy, S.E., Li, J.Z., Liu, C., Lucae, S., Maaser, A., MacIntyre, D.J., Mahon, P.B., Maier, W., Martinsson, L., McCarroll, S., McGuffin, P., McInnis, M.G., McKay, J.D., Medeiros, H., Medland, S.E., Meng, F., Milani, L., Montgomery, G.W., Morris, D.W., Mühleisen, T.W., Mullins, N., Nguyen, H., Nievergelt, C.M., Adolfsson, A.N., Nwulia, E.A., O’Donovan, C., Loohuis, L.M.O., Ori, A.P.S., Oruc, L., Ösby, U., Perlis, R.H., Perry, A., Pfennig, A., Potash, J.B., Purcell, S.M., Regeer, E.J., Reif, A., Reinbold, C.S., Rice, J.P., Rivas, F., Rivera, M., Roussos, P., Ruderfer, D.M., Ryu, E., Sánchez-Mora, C., Schatzberg, A.F., Scheftner, W.A., Schork, N.J., Shannon Weickert, C., Shehktman, T., Shilling, P.D., Sigurdsson, E., Slaney, C., Smeland, O.B., Sobell, J.L., Søholm Hansen, C., Spijker, A.T., St Clair, D., Steffens, M., Strauss, J.S., Streit, F., Strohmaier, J., Szelinger, S., Thompson, R.C., Thorgeirsson, T.E., Treutlein, J., Vedder, H., Wang, W., Watson, S.J., Weickert, T.W., Witt, S.H., Xi, S., Xu, W., Young, A.H., Zandi, P., Zhang, P., Zöllner, S., Adolfsson, R., Agartz, I., Alda, M., Backlund, L., Baune, B.T., Bellivier, F., Berrettini, W.H., Biernacka, J.M., Blackwood, D.H.R., Boehnke, M., Børglum, A.D., Corvin, A., Craddock, N., Daly, M.J., Dannlowski, U., Esko, T., Etain, B., Frye, M., Fullerton, J.M., Gershon, E.S., Gill, M., Goes, F., Grigoroiu-Serbanescu, M., Hauser, J., Hougaard, D.M., Hultman, C.M., Jones, I., Jones, L.A., Kahn, R.S., Kirov, G., Landén, M., Leboyer, M., Lewis, C.M., Li, Q.S., Lissowska, J., Martin, N.G., Mayoral, F., McElroy, S.L., McIntosh, A.M., McMahon, F.J., Melle, I., Metspalu, A., Mitchell, P.B., Morken, G., Mors, O., Mortensen, P.B., Müller-Myhsok, B., Myers, R.M., Neale, B.M., Nimgaonkar, V., Nordentoft, M., Nöthen, M.M., O’Donovan, M.C., Oedegaard, K.J., Owen, M.J., Paciga, S.A., Pato, C., Pato, M.T., Posthuma, D., Ramos-Quiroga, J.A., Ribasés, M., Rietschel, M., Rouleau, G.A., Schalling, M., Schofield, P.R., Schulze, T.G., Serretti, A., Smoller, J.W., Stefansson, H., Stefansson, K., Stordal, E., Sullivan, P.F., Turecki, G., Vaaler, A.E., Vieta, E., Vincent, J.B., Werge, T., Nurnberger, J.I., Wray, N.R., Di Florio, A., Edenberg, H.J., Cichon, S., Ophoff, R.A., Scott, L.J., Andreassen, O.A., Kelsoe, J., Sklar, P., 2019. Genome-wide association study identifies 30 loci associated with bipolar disorder. Nat. Genet. 51, 793–803. https://doi.org/10.1038/s41588-019-0397-8

Strike, L.T., Hansell, N.K., Couvy-Duchesne, B., Thompson, P.M., de Zubicaray, G.I., McMahon, K.L., Wright, M.J., 2019. Genetic Complexity of Cortical Structure: Differences in Genetic and Environmental Factors Influencing Cortical Surface Area and Thickness. Cereb. Cortex N. Y. N 1991 29, 952–962. https://doi.org/10.1093/cercor/bhy002

Sydnor, V.J., Larsen, B., Bassett, D.S., Alexander-Bloch, A., Fair, D.A., Liston, C., Mackey, A.P., Milham, M.P., Pines, A., Roalf, D.R., Seidlitz, J., Xu, T., Raznahan, A., Satterthwaite, T.D., 2021. Neurodevelopment of the association cortices: Patterns, mechanisms, and implications for psychopathology. Neuron S0896627321004578. https://doi.org/10.1016/j.neuron.2021.06.016

Taquet, M., Smith, S.M., Prohl, A.K., Peters, J.M., Warfield, S.K., Scherrer, B., Harrison, P.J., 2021. A structural brain network of genetic vulnerability to psychiatric illness. Mol. Psychiatry 26, 2089–2100. https://doi.org/10.1038/s41380-020-0723-7

Teeuw, J., Brouwer, R.M., Koenis, M.M.G., Swagerman, S.C., Boomsma, D.I., Hulshoff Pol, H.E., 2019. Genetic Influences on the Development of Cerebral Cortical Thickness During Childhood and Adolescence in a Dutch Longitudinal Twin Sample: The Brainscale Study. Cereb. Cortex N. Y. N 1991 29, 978–993. https://doi.org/10.1093/cercor/bhy005

Thapar, A., Riglin, L., 2020. The importance of a developmental perspective in Psychiatry: what do recent genetic-epidemiological findings show? Mol. Psychiatry 1–9. https://doi.org/10.1038/s41380-020-0648-1

The Autism Spectrum Disorders Working Group of The Psychiatric Genomics Consortium, 2017. Meta-analysis of GWAS of over 16,000 individuals with autism spectrum disorder highlights a novel locus at 10q24.32 and a significant overlap with schizophrenia. Mol. Autism 8, 21. https://doi.org/10.1186/s13229-017-0137-9

Trubetskoy, V., Pardiñas, A.F., Qi, T., Panagiotaropoulou, G., Awasthi, S., Bigdeli, T.B., Bryois, J., Chen, C.-Y., Dennison, C.A., Hall, L.S., Lam, M., Watanabe, K., Frei, O., Ge, T., Harwood, J.C., Koopmans, F., Magnusson, S., Richards, A.L., Sidorenko, J., Wu, Y., Zeng, J., Grove, J., Kim, M., Li, Z., Voloudakis, G., Zhang, W., Adams, M., Agartz, I., Atkinson, E.G., Agerbo, E., Al Eissa, M., Albus, M., Alexander, M., Alizadeh, B.Z., Alptekin, K., Als, T.D., Amin, F., Arolt, V., Arrojo, M., Athanasiu, L., Azevedo, M.H., Bacanu, S.A., Bass, N.J., Begemann, M., Belliveau, R.A., Bene, J., Benyamin, B., Bergen, S.E., Blasi, G., Bobes, J., Bonassi, S., Braun, A., Bressan, R.A., Bromet, E.J., Bruggeman, R., Buckley, P.F., Buckner, R.L., Bybjerg-Grauholm, J., Cahn, W., Cairns, M.J., Calkins, M.E., Carr, V.J., Castle, D., Catts, S.V., Chambert, K.D., Chan, R.C.K., Chaumette, B., Cheng, W., Cheung, E.F.C., Chong, S.A., Cohen, D., Consoli, A., Cordeiro, Q., Costas, J., Curtis, C., Davidson, M., Davis, K.L., de Haan, L., Degenhardt, F., DeLisi, L.E., Demontis, D., Dickerson, F., Dikeos, D., Dinan, T., Djurovic, S., Duan, J., Ducci, G., Dudbridge, F., Eriksson, J.G., Fañanás, L., Faraone, S.V., Fiorentino, A., Forstner, A., Frank, J., Freimer, N.B., Fromer, M., Frustaci, A., Gadelha, A., Genovese, G., Gershon, E.S., Giannitelli, M., Giegling, I., Giusti-Rodríguez, P., Godard, S., Goldstein, J.I., González Peñas, J., González-Pinto, A., Gopal, S., Gratten, J., Green, M.F., Greenwood, T.A., Guillin, O., Gülöksüz, S., Gur, R.E., Gur, R.C., Gutiérrez, B., Hahn, E., Hakonarson, H., Haroutunian, V., Hartmann, A.M., Harvey, C., Hayward, C., Henskens, F.A., Herms, S., Hoffmann, P., Howrigan, D.P., Ikeda, M., Iyegbe, C., Joa, I., Julià, A., Kähler, A.K., Kam-Thong, T., Kamatani, Y., Karachanak-Yankova, S., Kebir, O., Keller, M.C., Kelly, B.J., Khrunin, A., Kim, S.-W., Klovins, J., Kondratiev, N., Konte, B., Kraft, J., Kubo, M., Kučinskas, V., Kučinskiene, Z.A., Kusumawardhani, A., Kuzelova-Ptackova, H., Landi, S., Lazzeroni, L.C., Lee, P.H., Legge, S.E., Lehrer, D.S., Lencer, R., Lerer, B., Li, Miaoxin, Lieberman, J., Light, G.A., Limborska, S., Liu, C.-M., Lönnqvist, J., Loughland, C.M., Lubinski, J., Luykx, J.J., Lynham, A., Macek, M., Mackinnon, A., Magnusson, P.K.E., Maher, B.S., Maier, W., Malaspina, D., Mallet, J., Marder, S.R., Marsal, S., Martin, A.R., Martorell, L., Mattheisen, M., McCarley, R.W., McDonald, C., McGrath, J.J., Medeiros, H., Meier, S., Melegh, B., Melle, I., Mesholam-Gately, R.I., Metspalu, A., Michie, P.T., Milani, L., Milanova, V., Mitjans, M., Molden, E., Molina, E., Molto, M.D., Mondelli, V., Moreno, C., Morley, C.P., Muntané, G., Murphy, K.C., Myin-Germeys, I., Nenadić, I., Nestadt, G., Nikitina-Zake, L., Noto, C., Nuechterlein, K.H., O’Brien, N.L., O’Neill, F.A., Oh, S.-Y., Olincy, A., Ota, V.K., Pantelis, C., Papadimitriou, G.N., Parellada, M., Paunio, T., Pellegrino, R., Periyasamy, S., Perkins, D.O., Pfuhlmann, B., Pietiläinen, O., Pimm, J., Porteous, D., Powell, J., Quattrone, D., Quested, D., Radant, A.D., Rampino, A., Rapaport, M.H., Rautanen, A., Reichenberg, A., Roe, C., Roffman, J.L., Roth, J., Rothermundt, M., Rutten, B.P.F., Saker-Delye, S., Salomaa, V., Sanjuan, J., Santoro, M.L., Savitz, A., Schall, U., Scott, R.J., Seidman, L.J., Sharp, S.I., Shi, J., Siever, L.J., Sigurdsson, E., Sim, K., Skarabis, N., Slominsky, P., So, H.-C., Sobell, J.L., Söderman, E., Stain, H.J., Steen, N.E., Steixner-Kumar, A.A., Stögmann, E., Stone, W.S., Straub, R.E., Streit, F., Strengman, E., Stroup, T.S., Subramaniam, M., Sugar, C.A., Suvisaari, J., Svrakic, D.M., Swerdlow, N.R., Szatkiewicz, J.P., Ta, T.M.T., Takahashi, A., Terao, C., Thibaut, F., Toncheva, D., Tooney, P.A., Torretta, S., Tosato, S., Tura, G.B., Turetsky, B.I., Üçok, A., Vaaler, A., van Amelsvoort, T., van Winkel, R., Veijola, J., Waddington, J., Walter, H., Waterreus, A., Webb, B.T., Weiser, M., Williams, N.M., Witt, S.H., Wormley, B.K., Wu, J.Q., Xu, Z., Yolken, R., Zai, C.C., Zhou, W., Zhu, F., Zimprich, F., Atbaşoğlu, E.C., Ayub, M., Benner, C., Bertolino, A., Black, D.W., Bray, N.J., Breen, G., Buccola, N.G., Byerley, W.F., Chen, W.J., Cloninger, C.R., Crespo-Facorro, B., Donohoe, G., Freedman, R., Galletly, C., Gandal, M.J., Gennarelli, M., Hougaard, D.M., Hwu, H.-G., Jablensky, A.V., McCarroll, S.A., Moran, J.L., Mors, O., Mortensen, P.B., Müller-Myhsok, B., Neil, A.L., Nordentoft, M., Pato, M.T., Petryshen, T.L., Pirinen, M., Pulver, A.E., Schulze, T.G., Silverman, J.M., Smoller, J.W., Stahl, E.A., Tsuang, D.W., Vilella, E., Wang, S.-H., Xu, S., Indonesia Schizophrenia Consortium, Dai, N., Wenwen, Q., Wildenauer, D. B., Agiananda, F., Amir, N., Antoni, R., Arsianti, T., Asmarahadi, A., Diatri, H., Djatmiko, P., Irmansyah, I., Khalimah, S., Kusumadewi, I., Kusumaningrum, P., Lukman, P.R., Nasrun, M.W., Safyuni, N.S., Prasetyawan, P., Semen, G., Siste, K., Tobing, H., Widiasih, N., Wiguna, T., Wulandari, D., Evalina, N., Hananto, A.J., Ismoyo, J.H., Marini, T.M., Henuhili, S., Reza, M., Yusnadewi, S., PsychENCODE Abyzov, A., Akbarian, S., Ashley-Koch, A., van Bakel, H., Breen, M., Brown, M., Bryois, J., Carlyle, B., Charney, A., Coetzee, G., Crawford, G., Dracheva, S., Emani, P., Farnham, P., Fromer, M., Galeev, T., Gandal, M., Gerstein, M., Giase, G., Girdhar, K., Goes, F., Grennan, K., Gu, M., Guerra, B., Gursoy, G., Hoffman, G., Hyde, T., Jaffe, A., Jiang, S., Jiang, Y., Kefi, A., Kim, Y., Kitchen, R., Knowles, J.A., Lay, F., Lee, D., Li, Mingfeng, Liu, C., Liu, S., Mattei, E., Navarro, F., Pan, X., Peters, M.A., Pinto, D., Pochareddy, S., Polioudakis, D., Purcaro, M., Purcell, S., Pratt, H., Reddy, T., Rhie, S., Roussos, Panagiotis, Rozowsky, J., Sanders, S., Sestan, N., Sethi, A., Shi, X., Shieh, A., Swarup, V., Szekely, A., Wang, D., Warrell, J., Weissman, S., Weng, Z., White, K., Wiseman, J., Witt, H., Won, H., Wood, S., Wu, F., Xu, X., Yao, L., Zandi, P., Psychosis Endophenotypes International Consortium, Arranz, M.J., Bakker, S., Bender, S., Bramon, E., Collier, D.A., Crepo-Facorro, B., Hall, J., Iyegbe, C., Kahn, R., Lawrie, S., Lewis, C., Lin, K., Linszen, D.H., Mata, I., McIntosh, A., Murray, R.M., Ophoff, R.A., van Os, J., Powell, J., Rujescu, D., Walshe, M., Weisbrod, M., The SynGO Consortium, Achsel, T., Andres-Alonso, M., Bagni, C., Bayés, À., Biederer, T., Brose, N., Brown, T.C., Chua, J.J.E., Coba, M.P., Cornelisse, L.N., de Jong, A.P.H., de Juan-Sanz, J., Dieterich, D.C., Feng, G., Goldschmidt, H.L., Gundelfinger, E.D., Hoogenraad, C., Huganir, R.L., Hyman, S.E., Imig, C., Jahn, R., Jung, H., Kaeser, P.S., Kim, E., Koopmans, F., Kreutz, M.R., Lipstein, N., MacGillavry, H.D., Malenka, R., McPherson, P.S., O’Connor, V., Pielot, R., Ryan, T.A., Sahasrabudhe, D., Sala, C., Sheng, M., Smalla, K.-H., Smit, A.B., Südhof, T.C., Thomas, P.D., Toonen, R.F., van Weering, J.R.T., Verhage, M., Verpelli, C., Adolfsson, R., Arango, C., Baune, B.T., Belangero, S.I., Børglum, A.D., Braff, D., Bramon, E., Buxbaum, J.D., Campion, D., Cervilla, J.A., Cichon, S., Collier, D.A., Corvin, A., Curtis, D., Forti, M.D., Domenici, E., Ehrenreich, H., Escott-Price, V., Esko, T., Fanous, A.H., Gareeva, A., Gawlik, M., Gejman, P.V., Gill, M., Glatt, S.J., Golimbet, V., Hong, K.S., Hultman, C.M., Hyman, S.E., Iwata, N., Jönsson, E.G., Kahn, R.S., Kennedy, J.L., Khusnutdinova, E., Kirov, G., Knowles, J.A., Krebs, M.-O., Laurent-Levinson, C., Lee, J., Lencz, T., Levinson, D.F., Li, Q.S., Liu, J., Malhotra, A.K., Malhotra, D., McIntosh, A., McQuillin, A., Menezes, P.R., Morgan, V.A., Morris, D.W., Mowry, B.J., Murray, R.M., Nimgaonkar, V., Nöthen, M.M., Ophoff, R.A., Paciga, S.A., Palotie, A., Pato, C.N., Qin, S., Rietschel, M., Riley, B.P., Rivera, M., Rujescu, D., Saka, M.C., Sanders, A.R., Schwab, S.G., Serretti, A., Sham, P.C., Shi, Y., St Clair, D., Stefánsson, H., Stefansson, K., Tsuang, M.T., van Os, J., Vawter, M.P., Weinberger, D.R., Werge, T., Wildenauer, Dieter B., Yu, X., Yue, W., Holmans, P.A., Pocklington, A.J., Roussos, Panos, Vassos, E., Verhage, M., Visscher, P.M., Yang, J., Posthuma, D., Andreassen, O.A., Kendler, K.S., Owen, M.J., Wray, N.R., Daly, M.J., Huang, H., Neale, B.M., Sullivan, P.F., Ripke, S., Walters, J.T.R., O’Donovan, M.C., Schizophrenia Working Group of the Psychiatric Genomics Consortium, de Haan, L., van Amelsvoort, T., van Winkel, R., Gareeva, A., Sham, P.C., Shi, Y., St Clair, D., van Os, J., 2022. Mapping genomic loci implicates genes and synaptic biology in schizophrenia. Nature 604, 502–508. https://doi.org/10.1038/s41586-022-04434-5

Valk, S.L., Xu, T., Margulies, D.S., Masouleh, S.K., Paquola, C., Goulas, A., Kochunov, P., Smallwood, J., Yeo, B.T.T., Bernhardt, B.C., Eickhoff, S.B., 2020. Shaping brain structure: Genetic and phylogenetic axes of macroscale organization of cortical thickness. Sci. Adv. 6, eabb3417. https://doi.org/10.1126/sciadv.abb3417

van der Meer, Dennis, for the Pediatric Imaging, Neurocognition and Genetics Study, van der Meer, D., Rokicki, J., Kaufmann, T., Córdova-Palomera, A., Moberget, T., Alnæs, D., Bettella, F., Frei, O., Doan, N.T., Sønderby, I.E., Smeland, O.B., Agartz, I., Bertolino, A., Bralten, J., Brandt, C.L., Buitelaar, J.K., Djurovic, S., van Donkelaar, M., Dørum, E.S., Espeseth, T., Faraone, S.V., Fernández, G., Fisher, S.E., Franke, B., Haatveit, B., Hartman, C.A., Hoekstra, P.J., Håberg, A.K., Jönsson, E.G., Kolskår, K.K., Le Hellard, S., Lund, M.J., Lundervold, A.J., Lundervold, A., Melle, I., Monereo Sánchez, J., Norbom, L.C., Nordvik, J.E., Nyberg, L., Oosterlaan, J., Papalino, M., Papassotiropoulos, A., Pergola, G., de Quervain, D.J.F., Richard, G., Sanders, A.-M., Selvaggi, P., Shumskaya, E., Steen, V.M., Tønnesen, S., Ulrichsen, K.M., Zwiers, M.P., Andreassen, O.A., Westlye, L.T., 2020. Brain scans from 21,297 individuals reveal the genetic architecture of hippocampal subfield volumes. Mol. Psychiatry 25, 3053–3065. https://doi.org/10.1038/s41380-018-0262-7

van Erp, T.G.M., Walton, E., Hibar, D.P., Schmaal, L., Jiang, W., Glahn, D.C., Pearlson, G.D., Yao, N., Fukunaga, M., Hashimoto, R., Okada, N., Yamamori, H., Bustillo, J.R., Clark, V.P., Agartz, I., Mueller, B.A., Cahn, W., de Zwarte, S.M.C., Hulshoff Pol, H.E., Kahn, R.S., Ophoff, R.A., van Haren, N.E.M., Andreassen, O.A., Dale, A.M., Doan, N.T., Gurholt, T.P., Hartberg, C.B., Haukvik, U.K., Jørgensen, K.N., Lagerberg, T.V., Melle, I., Westlye, L.T., Gruber, O., Kraemer, B., Richter, A., Zilles, D., Calhoun, V.D., Crespo-Facorro, B., Roiz-Santiañez, R., Tordesillas-Gutiérrez, D., Loughland, C., Carr, V.J., Catts, S., Cropley, V.L., Fullerton, J.M., Green, M.J., Henskens, F.A., Jablensky, A., Lenroot, R.K., Mowry, B.J., Michie, P.T., Pantelis, C., Quidé, Y., Schall, U., Scott, R.J., Cairns, M.J., Seal, M., Tooney, P.A., Rasser, P.E., Cooper, G., Shannon Weickert, C., Weickert, T.W., Morris, D.W., Hong, E., Kochunov, P., Beard, L.M., Gur, R.E., Gur, R.C., Satterthwaite, T.D., Wolf, D.H., Belger, A., Brown, G.G., Ford, J.M., Macciardi, F., Mathalon, D.H., O’Leary, D.S., Potkin, S.G., Preda, A., Voyvodic, J., Lim, K.O., McEwen, S., Yang, F., Tan, Y., Tan, S., Wang, Z., Fan, F., Chen, J., Xiang, H., Tang, S., Guo, H., Wan, P., Wei, D., Bockholt, H.J., Ehrlich, S., Wolthusen, R.P.F., King, M.D., Shoemaker, J.M., Sponheim, S.R., De Haan, L., Koenders, L., Machielsen, M.W., van Amelsvoort, T., Veltman, D.J., Assogna, F., Banaj, N., de Rossi, P., Iorio, M., Piras, F., Spalletta, G., McKenna, P.J., Pomarol-Clotet, E., Salvador, R., Corvin, A., Donohoe, G., Kelly, S., Whelan, C.D., Dickie, E.W., Rotenberg, D., Voineskos, A.N., Ciufolini, S., Radua, J., Dazzan, P., Murray, R., Reis Marques, T., Simmons, A., Borgwardt, S., Egloff, L., Harrisberger, F., Riecher-Rössler, A., Smieskova, R., Alpert, K.I., Wang, L., Jönsson, E.G., Koops, S., Sommer, I.E.C., Bertolino, A., Bonvino, A., Di Giorgio, A., Neilson, E., Mayer, A.R., Stephen, J.M., Kwon, J.S., Yun, J.-Y., Cannon, D.M., McDonald, C., Lebedeva, I., Tomyshev, A.S., Akhadov, T., Kaleda, V., Fatouros-Bergman, H., Flyckt, L., Busatto, G.F., Rosa, P.G.P., Serpa, M.H., Zanetti, M.V., Hoschl, C., Skoch, A., Spaniel, F., Tomecek, D., Hagenaars, S.P., McIntosh, A.M., Whalley, H.C., Lawrie, S.M., Knöchel, C., Oertel-Knöchel, V., Stäblein, M., Howells, F.M., Stein, D.J., Temmingh, H.S., Uhlmann, A., Lopez-Jaramillo, C., Dima, D., McMahon, A., Faskowitz, J.I., Gutman, B.A., Jahanshad, N., Thompson, P.M., Turner, J.A., Farde, L., Flyckt, L., Engberg, G., Erhardt, S., Fatouros-Bergman, H., Cervenka, S., Schwieler, L., Piehl, F., Agartz, I., Collste, K., Victorsson, P., Malmqvist, A., Hedberg, M., Orhan, F., 2018. Cortical Brain Abnormalities in 4474 Individuals With Schizophrenia and 5098 Control Subjects via the Enhancing Neuro Imaging Genetics Through Meta Analysis (ENIGMA) Consortium. Biol. Psychiatry 84, 644–654. https://doi.org/10.1016/j.biopsych.2018.04.023

Weinberger, D.R., 2017. The neurodevelopmental origins of schizophrenia in the penumbra of genomic medicine. World Psychiatry 16, 225–226. https://doi.org/10.1002/wps.20474

Weinberger, D.R., 1987. Implications of Normal Brain Development for the Pathogenesis of Schizophrenia. Arch. Gen. Psychiatry 44, 660. https://doi.org/10.1001/archpsyc.1987.01800190080012

Westlye, L.T., Alnæs, D., van der Meer, D., Kaufmann, T., Andreassen, O.A., 2019. Population-Based Mapping of Polygenic Risk for Schizophrenia on the Human Brain: New Opportunities to Capture the Dimensional Aspects of Severe Mental Disorders. Biol. Psychiatry 86, 499–501. https://doi.org/10.1016/j.biopsych.2019.08.001

Westlye, L.T., Walhovd, K.B., Dale, A.M., Bjørnerud, A., Due-Tønnessen, P., Engvig, A., Grydeland, H., Tamnes, C.K., Ostby, Y., Fjell, A.M., 2010. Life-span changes of the human brain white matter: diffusion tensor imaging (DTI) and volumetry. Cereb. Cortex N. Y. N 1991 20, 2055–2068. https://doi.org/10.1093/cercor/bhp280

White, N.S., Leergaard, T.B., D’Arceuil, H., Bjaalie, J.G., Dale, A.M., 2013. Probing tissue microstructure with restriction spectrum imaging: Histological and theoretical validation. Hum. Brain Mapp. 34, 327–346. https://doi.org/10.1002/hbm.21454

Wray, N.R., Ripke, S., Mattheisen, M., Trzaskowski, M., Byrne, E.M., Abdellaoui, A., Adams, M.J., Agerbo, E., Air, T.M., Andlauer, T.M.F., Bacanu, S.-A., Bækvad-Hansen, M., Beekman, A.F.T., Bigdeli, T.B., Binder, E.B., Blackwood, D.R.H., Bryois, J., Buttenschøn, H.N., Bybjerg-Grauholm, J., Cai, N., Castelao, E., Christensen, J.H., Clarke, T.-K., Coleman, J.I.R., Colodro-Conde, L., Couvy-Duchesne, B., Craddock, N., Crawford, G.E., Crowley, C.A., Dashti, H.S., Davies, G., Deary, I.J., Degenhardt, F., Derks, E.M., Direk, N., Dolan, C.V., Dunn, E.C., Eley, T.C., Eriksson, N., Escott-Price, V., Kiadeh, F.H.F., Finucane, H.K., Forstner, A.J., Frank, J., Gaspar, H.A., Gill, M., Giusti-Rodríguez, P., Goes, F.S., Gordon, S.D., Grove, J., Hall, L.S., Hannon, E., Hansen, C.S., Hansen, T.F., Herms, S., Hickie, I.B., Hoffmann, P., Homuth, G., Horn, C., Hottenga, J.-J., Hougaard, D.M., Hu, M., Hyde, C.L., Ising, M., Jansen, R., Jin, F., Jorgenson, E., Knowles, J.A., Kohane, I.S., Kraft, J., Kretzschmar, W.W., Krogh, J., Kutalik, Z., Lane, J.M., Li, Yihan, Li, Yun, Lind, P.A., Liu, X., Lu, L., MacIntyre, D.J., MacKinnon, D.F., Maier, R.M., Maier, W., Marchini, J., Mbarek, H., McGrath, P., McGuffin, P., Medland, S.E., Mehta, D., Middeldorp, C.M., Mihailov, E., Milaneschi, Y., Milani, L., Mill, J., Mondimore, F.M., Montgomery, G.W., Mostafavi, S., Mullins, N., Nauck, M., Ng, B., Nivard, M.G., Nyholt, D.R., O’Reilly, P.F., Oskarsson, H., Owen, M.J., Painter, J.N., Pedersen, C.B., Pedersen, M.G., Peterson, R.E., Pettersson, E., Peyrot, W.J., Pistis, G., Posthuma, D., Purcell, S.M., Quiroz, J.A., Qvist, P., Rice, J.P., Riley, B.P., Rivera, M., Saeed Mirza, S., Saxena, R., Schoevers, R., Schulte, E.C., Shen, L., Shi, J., Shyn, S.I., Sigurdsson, E., Sinnamon, G.B.C., Smit, J.H., Smith, D.J., Stefansson, H., Steinberg, S., Stockmeier, C.A., Streit, F., Strohmaier, J., Tansey, K.E., Teismann, H., Teumer, A., Thompson, W., Thomson, P.A., Thorgeirsson, T.E., Tian, C., Traylor, M., Treutlein, J., Trubetskoy, V., Uitterlinden, A.G., Umbricht, D., Van der Auwera, S., van Hemert, A.M., Viktorin, A., Visscher, P.M., Wang, Y., Webb, B.T., Weinsheimer, S.M., Wellmann, J., Willemsen, G., Witt, S.H., Wu, Y., Xi, H.S., Yang, J., Zhang, F., Arolt, V., Baune, B.T., Berger, K., Boomsma, D.I., Cichon, S., Dannlowski, U., de Geus, E.C.J., DePaulo, J.R., Domenici, E., Domschke, K., Esko, T., Grabe, H.J., Hamilton, S.P., Hayward, C., Heath, A.C., Hinds, D.A., Kendler, K.S., Kloiber, S., Lewis, G., Li, Q.S., Lucae, S., Madden, P.F.A., Magnusson, P.K., Martin, N.G., McIntosh, A.M., Metspalu, A., Mors, O., Mortensen, P.B., Müller-Myhsok, B., Nordentoft, M., Nöthen, M.M., O’Donovan, M.C., Paciga, S.A., Pedersen, N.L., Penninx, B.W.J.H., Perlis, R.H., Porteous, D.J., Potash, J.B., Preisig, M., Rietschel, M., Schaefer, C., Schulze, T.G., Smoller, J.W., Stefansson, K., Tiemeier, H., Uher, R., Völzke, H., Weissman, M.M., Werge, T., Winslow, A.R., Lewis, C.M., Levinson, D.F., Breen, G., Børglum, A.D., Sullivan, P.F., 2018. Genome-wide association analyses identify 44 risk variants and refine the genetic architecture of major depression. Nat. Genet. 50, 668–681. https://doi.org/10.1038/s41588-018-0090-3

Yang, J., Benyamin, B., McEvoy, B.P., Gordon, S., Henders, A.K., Nyholt, D.R., Madden, P.A., Heath, A.C., Martin, N.G., Montgomery, G.W., Goddard, M.E., Visscher, P.M., 2010. Common SNPs explain a large proportion of the heritability for human height. Nat. Genet. 42, 565–569. https://doi.org/10.1038/ng.608

Yang, J., Lee, S.H., Goddard, M.E., Visscher, P.M., 2011. GCTA: a tool for genome-wide complex trait analysis. Am. J. Hum. Genet. 88, 76–82. https://doi.org/10.1016/j.ajhg.2010.11.011

Yilmaz, M., Yalcin, E., Presumey, J., Aw, E., Ma, M., Whelan, C.W., Stevens, B., McCarroll, S.A., Carroll, M.C., 2021. Overexpression of schizophrenia susceptibility factor human complement C4A promotes excessive synaptic loss and behavioral changes in mice. Nat. Neurosci. 24, 214–224. https://doi.org/10.1038/s41593-020-00763-8

Zhao, B., Li, T., Yang, Y., Wang, X., Luo, T., Shan, Y., Zhu, Z., Xiong, D., Hauberg, M.E., Bendl, J., Fullard, J.F., Roussos, P., Li, Y., Stein, J.L., Zhu, H., 2021. Common genetic variation influencing human white matter microstructure. Science 372, eabf3736. https://doi.org/10.1126/science.abf3736

Zhou, D., Lebel, C., Treit, S., Evans, A., Beaulieu, C., 2015. Accelerated longitudinal cortical thinning in adolescence. NeuroImage 104, 138–145. https://doi.org/10.1016/j.neuroimage.2014.10.005

