## supplementary materials for "Associations between brain imaging and polygenic scores of mental health and educational attainment in children aged 9-11"

**Imputation quality checks**

We assessed the imputation quality of MRI and the behavioral data based on descriptive checks. The descriptive checks included a plot of the percentage of missing data per variable, the frequency of missingness combinations, and density distribution plots of the observed and imputed data. For the MRI data, panel A of the figure below shows that the maximum percentage of missing values per variable was slightly above 4% while most variables had less than 1% of missing data. Panel B shows the combinations of missing values per variable. The combination that was most frequent was the one with complete observations (~38%), while the second most frequent combinations (including missing data) were much less frequent (< 0.5%). This shows that no systematic patterns of missingness combinations were observed in the data. Panel C shows the distributions of the observed and imputed MRI data per imaging modality. Great overlap between the distributions was observed across modalities, showing that imputed data were plausible and in similar value ranges than the observed data. However, it is not be necessarily problematic that imputed values deviate from the observed data ranges (Nguyen et al., 2017). It is important to mention that these descriptive checks do not formally test for statistical differences between the distributions of observed and imputed data, so additional tests were performed to ensure the validity of our results (see Supplementary Figure 8). Similar observations of imputation quality are seen with the behavioral (CBCL) data. See the second figure below.

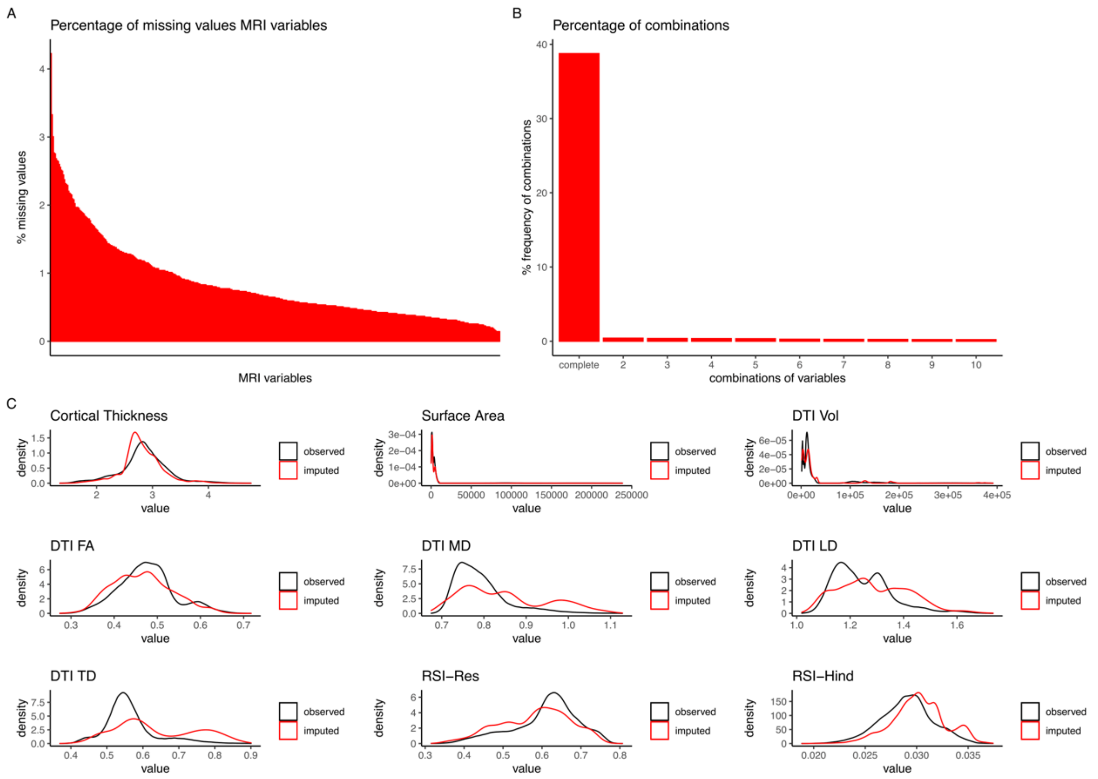
A
Percentage of missing values MRI variables
% missing values
MRI variables
B
Percentage of combinations
% frequency of combinations
combinations of variables
complete
C
Cortical Thickness
Surface Area
DTI Vol
DTI FA
DTI MD
DTI LD
DTI TD
RSI−Res
RSI−Hind
density
value
observed
imputed

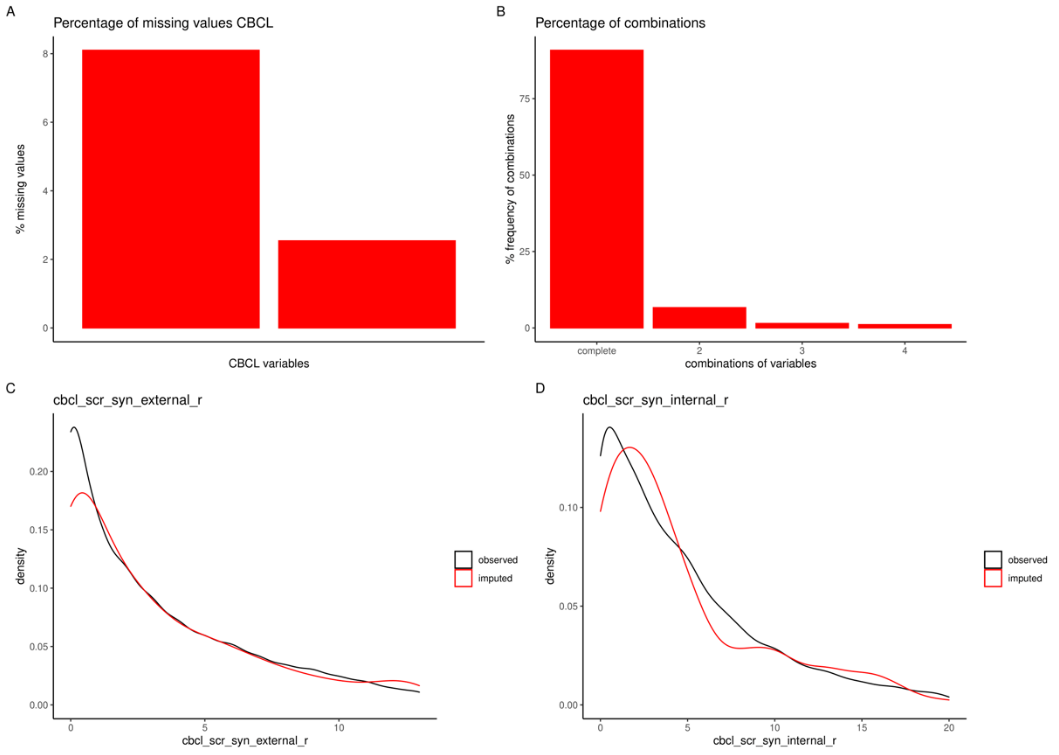
A
Percentage of missing values CBCL
% missing values
CBCL variables
B
Percentage of combinations
% frequency of combinations
combinations of variables
complete
C
cbcl_scr_syn_external_r
D
cbcl_scr_syn_internal_r
density
observed
imputed

**Supplementary Figure 1. PCA of the MRI modalities.** Scree plots showing each PC on the x axis and the variance explained on the y axis. The red lines mark the variance explained by the first three components. The blue lines show the cumulative variance explained.

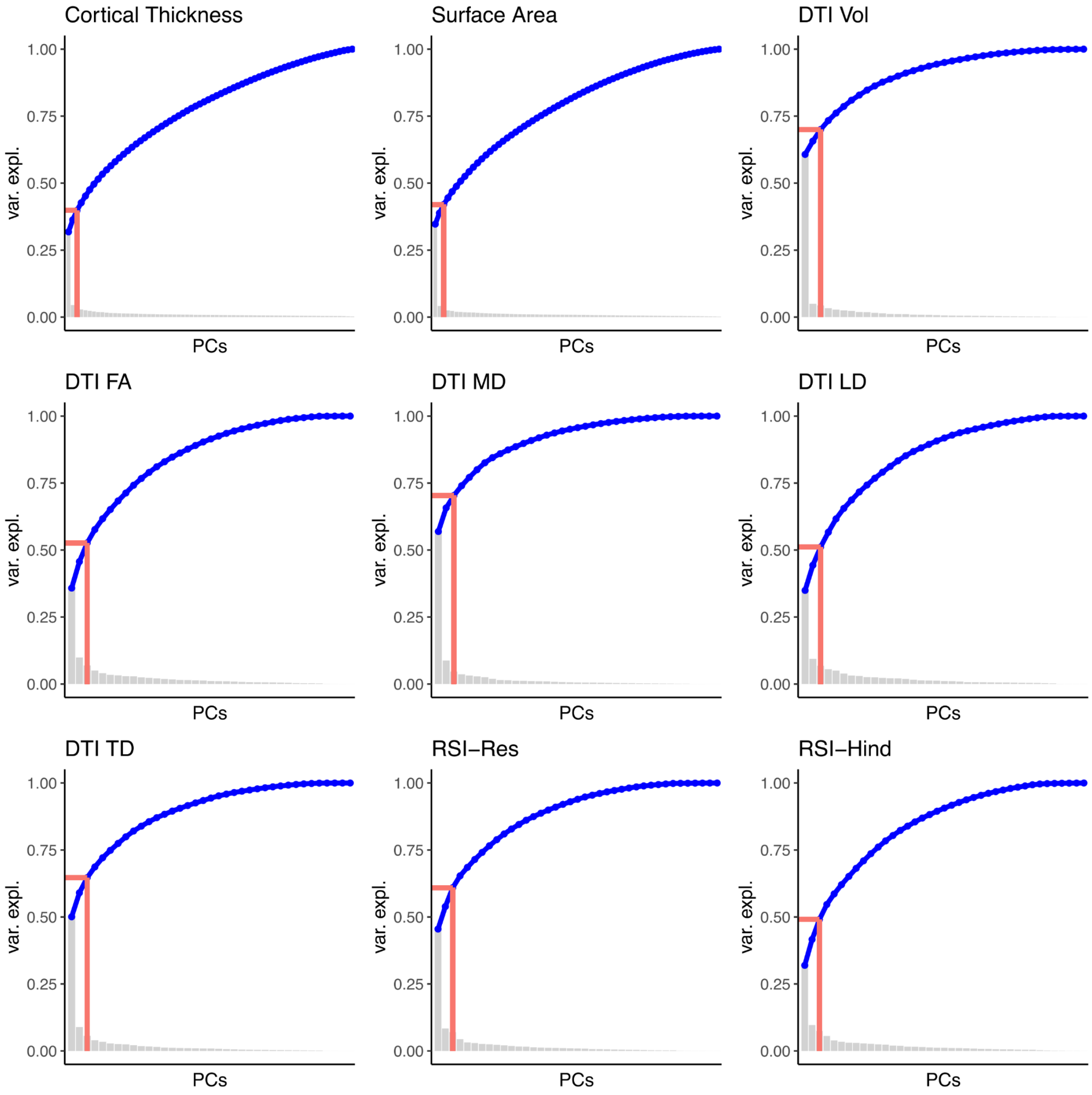

**Supplementary Figure 2. Labels diffusion plots.**

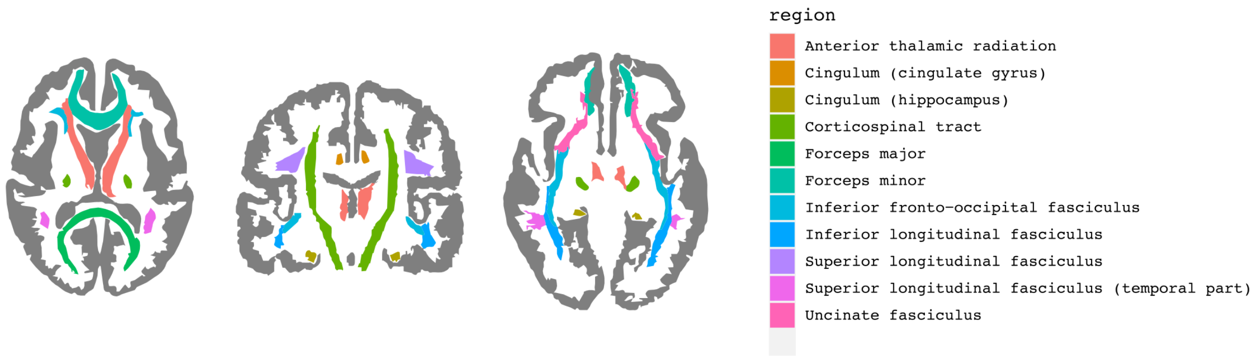

**Supplementary Figure 3. Principal component loadings for diffusion modalities.** Brain plots display the loadings of the variables on each of the first three components. Note that some tracts are not presented in these graphs (e.g., fornix, corpus callosum or corticostriatal tracts). Full loadings are reported in supplementary table S1.

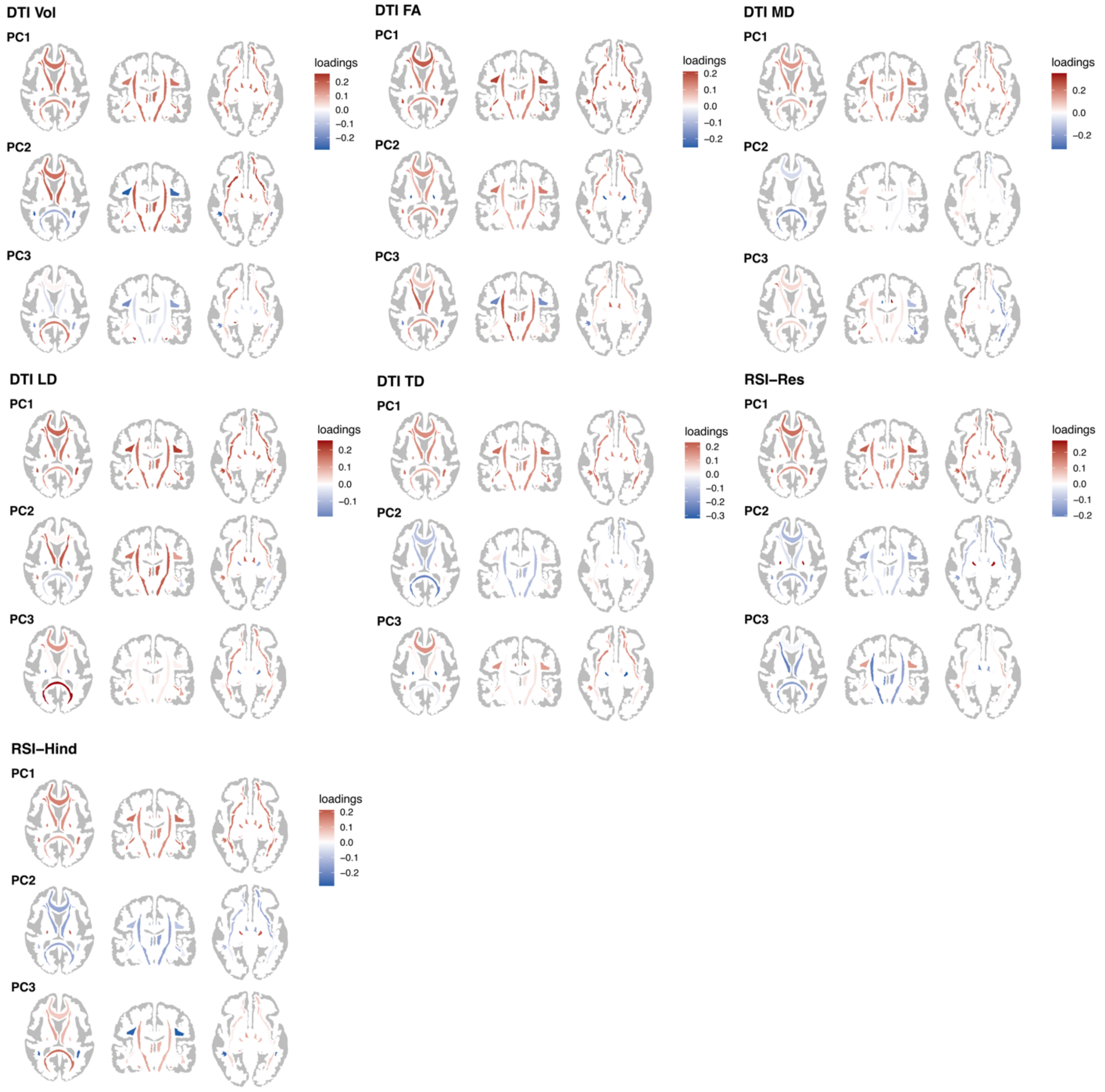

**Supplementary Figure 4. SNP heritability of the imaging components in a subset of individuals with genetic European ancestry (n=3,841).** Heritability estimates largely resemble those observed in the full mixed sample. For instance, the regional surface area of occipital regions (PC2) exhibits one of the highest heritability estimates.

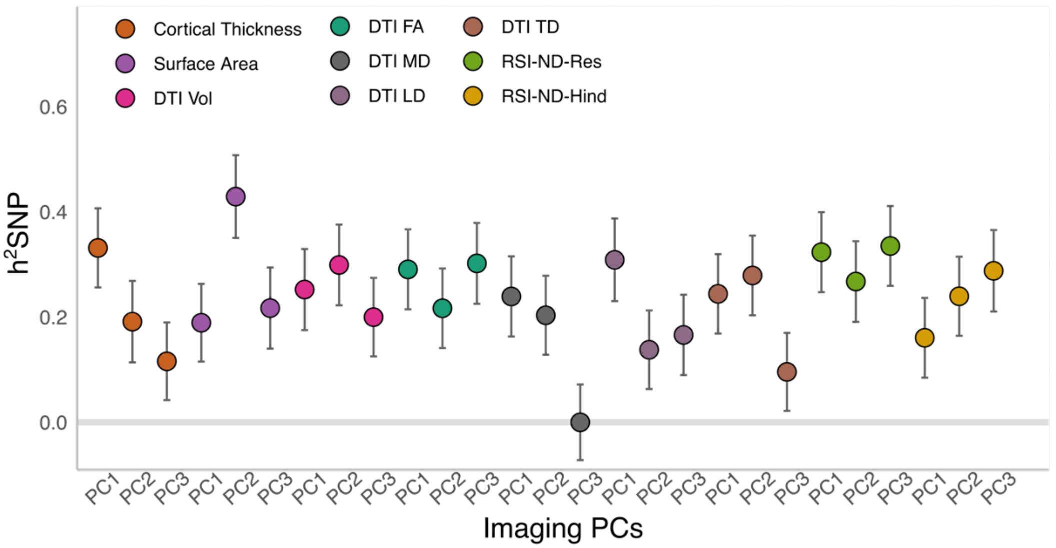

**Supplementary Figure 5. SNP heritability of the first 20 principal components in the full sample (n=7,124).** The dot plot shows the point estimates of the heritability of each of the first 20 brain imaging components of each modality. The dots are colored by imaging modality and displayed in order 1-20 on the x axis. The labels highlight the imaging components with the highest SNP heritability ($h^2SNP>0.25$). The y-axis shows the SNP heritability. Below the dotplot, brain plots display the first 20 principal component loadings for the cortical imaging modalities (red=positive loadings; blue=negative loadings).

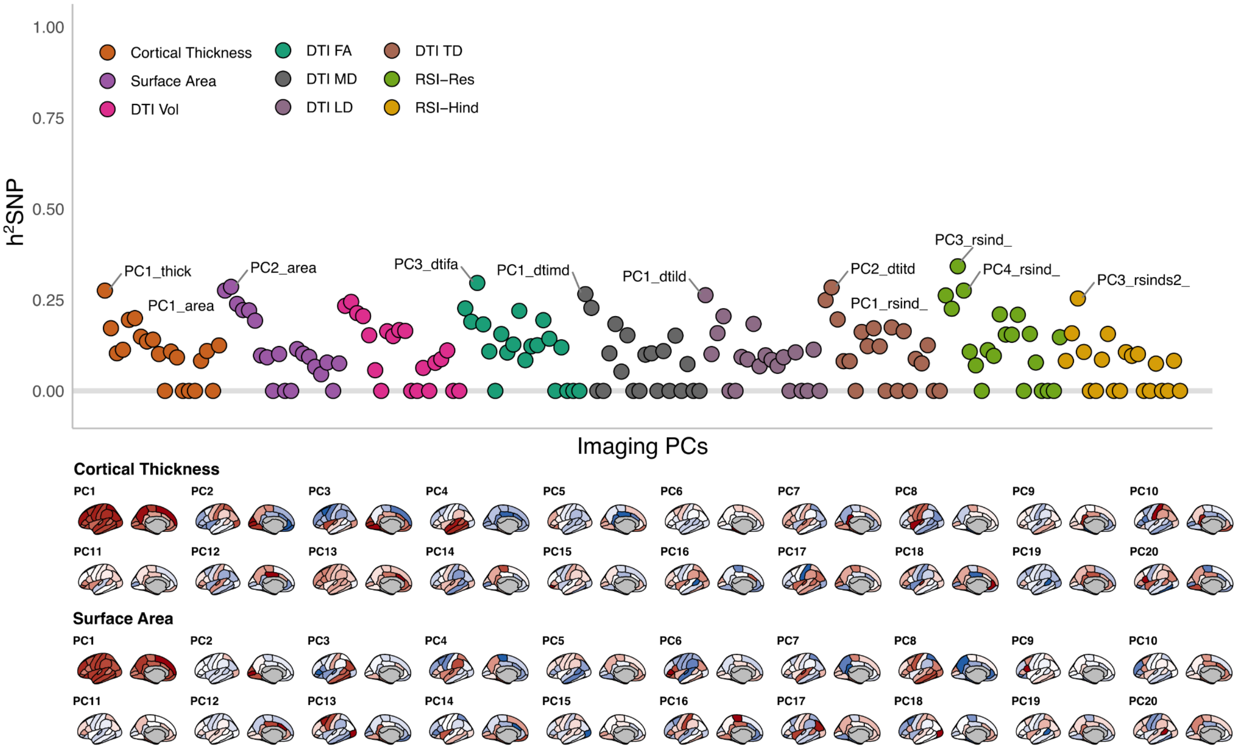

**Supplementary Figure 6. Posterior distributions of the univariate Bayesian models examining the relationships between PRS and imaging components.** Distributions of the posterior densities. Dots show the mean of the posterior distributions and lines represent the 95% Credible Interval.

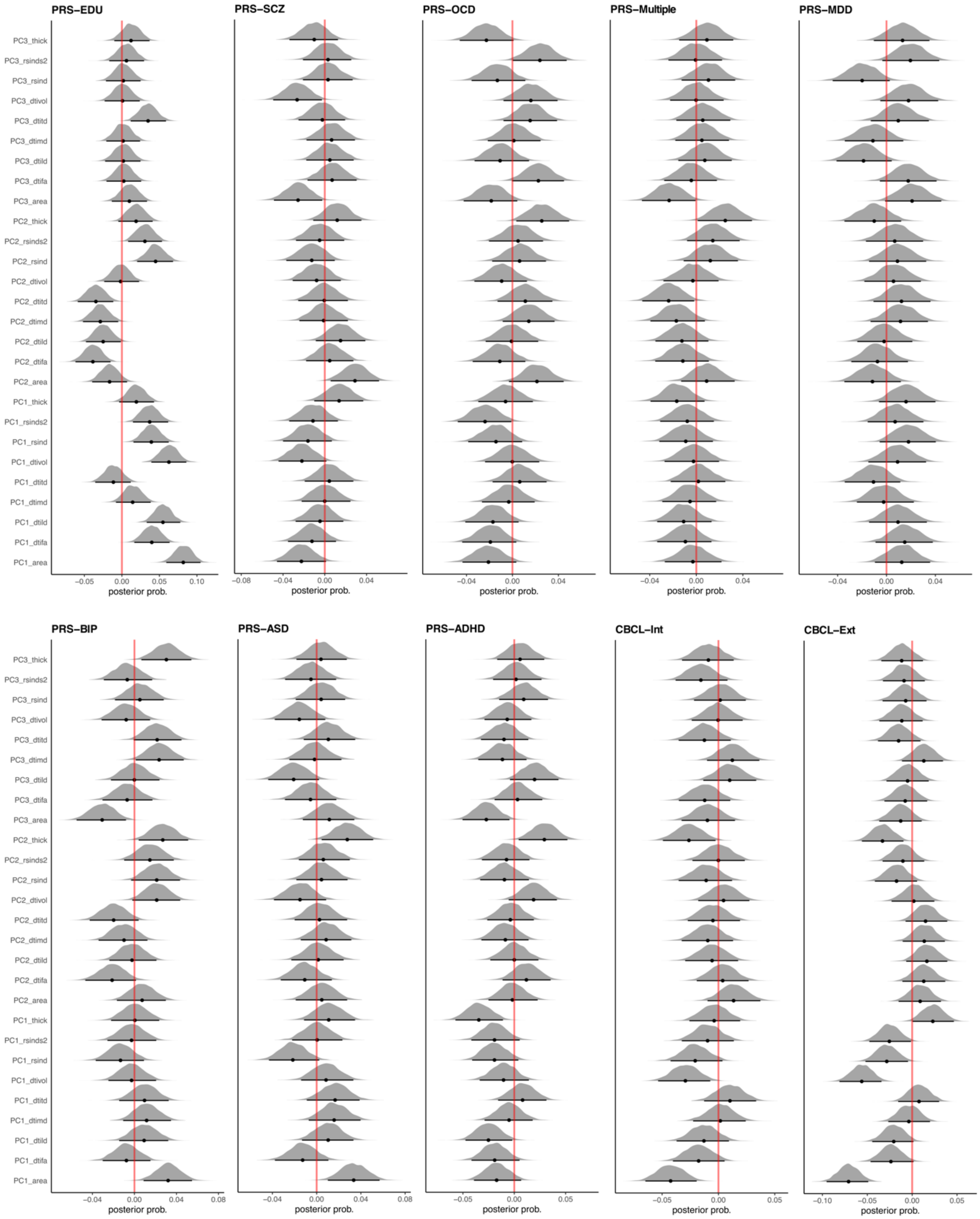

**Supplementary Figure 7. Univariate associations between brain imaging components and polygenic scores and clinical variables in a subset of individuals with genetic European ancestry (n=3,841).** On the left-hand side, posterior means of the univariate Bayesian linear regression models between brain imaging (x-axis) and PRS of psychiatric disorders and PRS-Edu and CBCL behavioral scores (y-axis) as comparison. On the right-hand side, Bayes factors showed largely null and anecdotal associations between psychiatric PRS and brain imaging components. As in the main sample, the strongest evidence showed associations between PRS-Edu, CBCL externalizing problems and global surface area and DTI volumes.

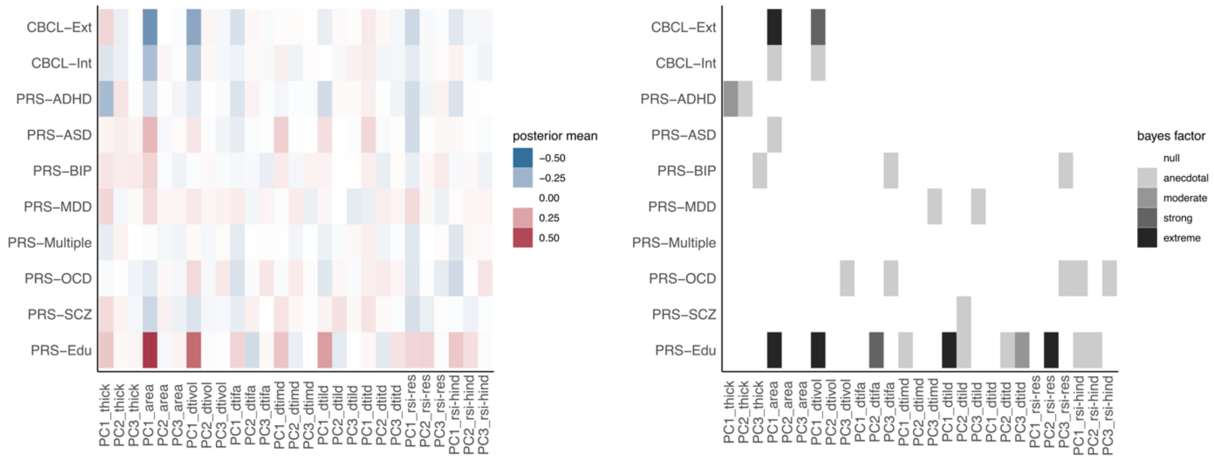

**Supplementary Figure 8. PCA and CCA in sample without excluding individuals based on extreme MRI data (n=7,334).** The results recapitulate the main findings excluding and imputing extreme observations. On the left-hand side**,** PCA loadings of the first three components of cortical thickness and surface area. On the right-hand side, CCA loadings of imaging components and PRS.

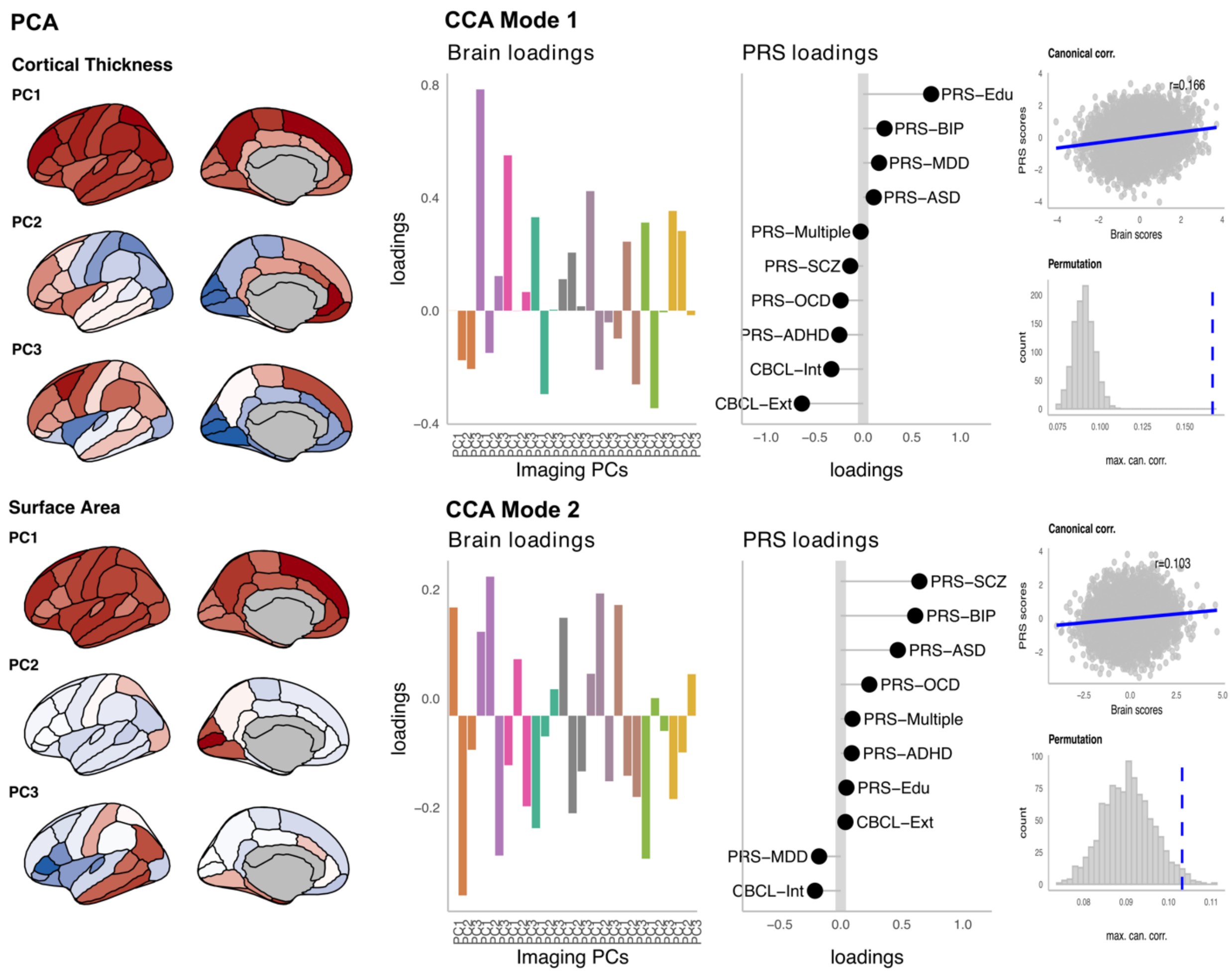

**Supplementary Figure 9. Brain scores predict fluid and crystallized cognition at baseline.** Posterior density distributions, showing the mean (dot) and 95% Credible Interval (black lines) of the posterior estimates. On the left-hand side, Mode 1 brain scores were associated with both fluid and crystalized NIH scores at baseline, whereas brain scores of Mode 2 were marginally associated with crystalized scores. On the right-hand side, estimates of the interaction between brain scores and Time show null effects of brain scores and changes in cognition over time.

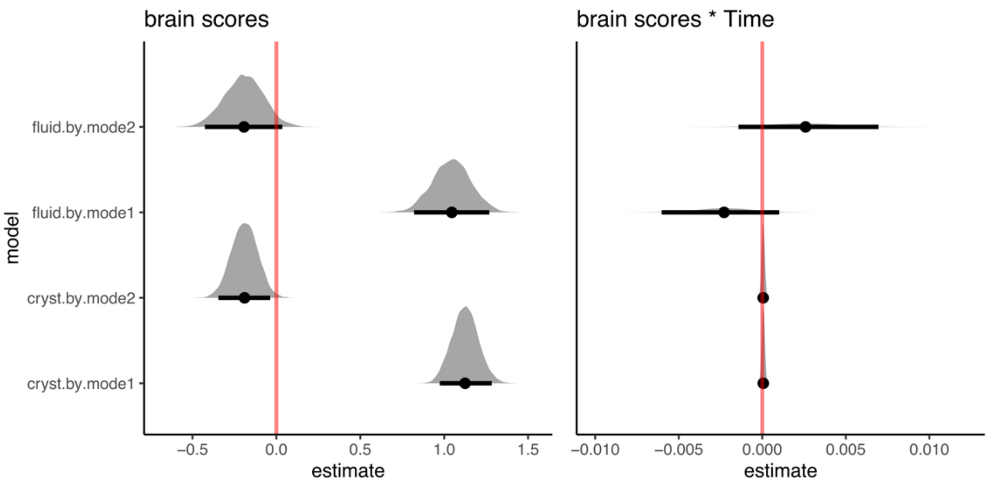

**Supplementary Figure 10. Chart of analyses steps.**

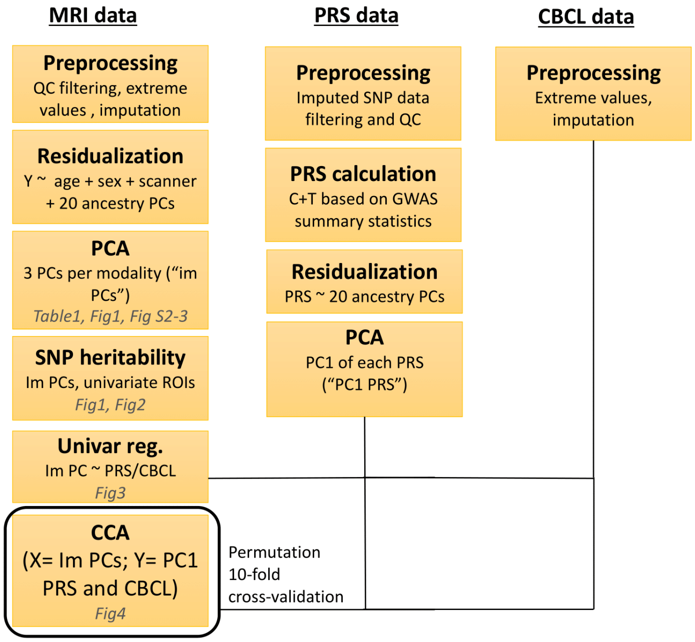
